# The nutritional and metabolic impact of night shift work in a real-world setting

**DOI:** 10.1101/2025.10.19.25338338

**Authors:** Friedrich C. Jassil, Nicholas E. Phillips, Alexandra Hemmer, Céline Joris, Andrew D. Biancolin, Steffen L. Hartmeyer, Victor Dorribo, Stephen Perrig, Laurence Genton, Virginie Sterpenich, Marcel Salathé, Jacques A. Pralong, Marilyne Andersen, Claude Pichard, Charna Dibner, Tinh-Hai Collet

## Abstract

**Background:** Growing evidence from large epidemiological and small lab-controlled studies links night shift work to impaired sleep, cognitive performance, and cardiometabolic health, but evidence from real-world occupational environments remains limited.

**Methods:** Nutritional intake and cardiometabolic parameters were assessed using multiple wearable devices and mobile health apps, alongside other objective and subjective assessment tools among shift workers during three consecutive night shifts and three consecutive day shifts.

**Results:** Of 72 participants, 96% were healthcare workers, 82% were female, with a median age of 31 years and a median body mass index 22.7 Kg/m^2^. Night shifts led to poorer cognitive performance and sleep quality with higher levels of fatigue (all p<0.01). While intake of calories, protein, and fat was reduced during night shifts (all p<0.05), glycemic variability and postprandial glucose spikes were increased compared to day shifts (all p<0.05). After three consecutive night shifts, the morning systolic blood pressure was elevated, plasma cortisol was reduced (both p<0.05), and the 24-hour heart rate and heart rate variability were altered, indicating disrupted parasympathetic function. Exploratory analysis revealed higher protein intake correlated with lower glycemic variability (p<0.01), whereas nighttime eating was associated with increased glycemic variability (p<0.05).

**Conclusion:** Night shift work is associated with adverse cardiometabolic health. Future interventional studies could target meal composition and meal timing to improve glycemic variability in night shift workers.

**Trial Registration:** ClinicalTrials.gov NCT05177965

## INTRODUCTION

The proportion of shift workers is increasing and now represents 20-25% of the global workforce (1). In Switzerland, approximately 700,000 employees are subjected to irregular working hours, such as rotating shifts (a series of day shifts, evening shifts, and night shifts, then rotating from one type of shift to the next) or permanent night shifts (2). These work patterns are common in sectors such as healthcare and emergency services, transportation, manufacturing, and retail services. Night shift work, associated with nocturnal light exposure, late meal consumption, and reduced sleep duration, can desynchronize endogenous body clocks from environmental cues, leading to circadian misalignment (3). Epidemiological evidence indicates that prolonged exposure to night shift work increases susceptibility to obesity, type 2 diabetes, hypertension, and metabolic syndrome, leading to long-term complications such as cancer and cardiovascular mortality (4).

Circadian misalignment protocols under lab-controlled conditions simulating shift work have provided important insights into the mechanisms underlying the negative consequences induced by night shift work (5–7). A 3-day simulated night shift work led to decreased insulin sensitivity and increased fasting plasma glucose levels (7). Similarly, an 8-day simulated night shift resulted in elevated postprandial plasma glucose, despite a concomitant increase in plasma insulin levels. Circadian misalignment also led to increased blood pressure and impaired attention, the latter being associated with increased sleepiness (5). Interestingly, the circadian misalignment protocol in healthy chronic shift workers resulted in impaired glucose tolerance and reduced insulin sensitivity. Circadian misalignment also led to reduced total sleep time and increased 24-hour blood pressure (6). However, it is important to note that the tightly controlled lab conditions do not fully replicate the various environmental and behavioral factors present in real-life settings of shift workers that can impact circadian rhythms. These factors include the variation in exposure to ambient light and temperature, the actual workload, physical and emotional exhaustion, variability in dietary and physical activity patterns, as well as smoking and caffeine intake. Therefore, data from observational studies within real-world occupational settings are needed to bridge the knowledge gap between epidemiological and lab-controlled experimental evidence (4).

Recent advances in wearable technology and mobile health (mHealth) applications (apps) have allowed for live data collection rather than retrospective self-reports, providing more accurate and real-time insights into health parameters and behaviors (8). These technologies include continuous glucose monitors (CGM) to assess glycemic profiles, triaxial actigraphy devices to measure physical activity, sleep-wake cycles, and heart rate (HR), ambient light sensors to track light exposure, body temperature sensors, and smartphone food diary apps to record food intake. Together, these tools provide continuous 24-hour insights into *in vivo* clock parameters during night shift work, addressing the limitations of earlier studies that relied on data collected from isolated time points. A deeper understanding of the physiological changes linked to lifestyle behaviors during night shift work may steer the development of targeted occupational health interventions to improve the health and well-being of night shift workers.

Using multiple wearable devices and mHealth apps, alongside other objective and subjective assessment tools, we aimed to evaluate the nutritional and metabolic health impacts of night shift work in a real-world environment in the OPTI-SHIFT observational study (**Supplemental Figure 1**, ClinicalTrials.gov registry no. NCT05177965). Other outcome data on light exposure and core body temperature from the same cohort have recently been published elsewhere (9).

## RESULTS

We enrolled 72 night shift workers, consisting of 96% of healthcare workers. Of all participants, 82% were female with a median age of 31 (IQR 28 to 35) years and a body mass index (BMI) of 22.7 (IQR 20.4 to 26.2) Kg/m^2^ (**Table 1 and Supplemental Table 1**). Two participants did not complete the series of day shifts due to withdrawal (*n* = 1) and loss of follow-up (*n* = 1). The self-reported shift schedules were between 21:40h ± 01:12h and 07:30h ± 00:42h for night shifts, and between 07:00h ± 00:36h and 16:30h ± 01:42h for day shifts. The median interval period between the series of night shifts (first time point) and the series of day shifts (second time point) was 10.3 (IQR 5.3 to 18.6) weeks. Participants reported an average of five years working in shift work, with three years in the current shift pattern averaging 40 hours per week. The detailed work patterns, shift work systems, motivations, and workload are provided in **Supplemental Table 2**.

**Table 1.** Participant characteristics. ^A^Sleep quality in the past month was assessed by the Pittsburgh Sleep Quality Index (PSQI), with a total score range of 1-21, higher scores indicating poorer sleep quality (63). See also Supplemental Table 1.

| **Participant characteristics** | ***n* = 72** |
| --- | --- |
| Age, years, median (IQR) | 31 (28; 35) |
| Women, *n* (%) | 59 (81.9) |
| Marital status, *n* (%) |  |
| Single | 37 (51.3) |
| Married, partnership, living with partner | 31 (43.1) |
| Separated, divorced | 4 (5.6) |
| **Employment and shift work** |  |
| Profession, *n* (%) |  |
| Nurse | 36 (50.0) |
| Physician | 20 (27.8) |
| Paramedic | 13 (18.1) |
| Others | 3 (4.1) |
| Duration of employment, years, median (IQR) | 6 (3.75; 11) |
| Duration of shift work employment | 5 (2.9; 10) |
| Duration of working in the present shift | 3 (1.3; 5.5) |
| Regularity of shift work system, *n* (%) |  |
| Regular | 8 (11.1) |
| Irregular | 57 (79.2) |
| Flexible | 7 (9.7) |
| **Clinical data** |  |
| Weight, Kg, median (IQR) | 64.4 (54.0; 75.1) |

**Table 1 legend:** <sup>A</sup>Sleep quality in the past month was assessed by the Pittsburgh Sleep Quality Index (PSQI), with a total score range of 1-21, higher scores indicating poorer sleep quality (63). See also Supplemental Table 1.
| Participant characteristics | *n* = 72 |
| --- | --- |
| Height, m, median (IQR) | 1.65 (1.60; 1.73) |
| Body mass index, Kg/m<sup>2</sup>, median (IQR) | 22.7 (20.4; 26.2) |
| Waist circumference, cm, median (IQR) | 77.4 (71.4; 86.1) |
| Waist-to-hip ratio, median (IQR) | 0.83 (0.78; 0.88) |
| Body composition by bioelectrical impedance analysis, median (IQR) |  |
| Fat mass (Kg) | 17.5 (14.2; 24.4) |
| Fat-free mass (Kg) | 43.3 (39.3; 48.9) |
| Fat mass index (Kg/m<sup>2</sup>) | 6.2 (5.2; 8.3) |
| Fat-free mass index (Kg/m<sup>2</sup>) | 15.9 (14.8; 17.4) |
| Sleep quality<sup>A</sup> (PSQI score, *n* = 69), mean ± SD | 7.72 ± 3.57 |

### Higher emotional stress but lighter time pressures were perceived during night shifts

In general, most participants stated that physical and mental workloads were about the same between night and day shifts. However, emotional stress was more often perceived as higher during night shifts, while time pressures were generally considered lighter during night shifts (**Supplemental Table 2**).

### Higher systolic and diastolic blood pressure, but lower heart rate and plasma cortisol in the morning following night shifts

The anthropometric and cardiometabolic parameters are compared between the two series of shifts in **Table 2**. We found no differences in body weight or BMI. However, higher morning systolic (+1.9 ± 6.9 mmHg, p=0.03) and diastolic blood pressure (+1.4 ± 6.0 mmHg, p-value close to borderline significance, p=0.052), but lower HR (-3.5 ± 10.0 beats/min, p=0.005) and plasma cortisol (-108 nmol/L, IQR -210 to 33, p=0.007) were observed post-night shift compared to the morning of day shift. There were no significant differences in other cardiometabolic parameters between the two series of shifts. Levels of self-reported physical activity, mental health, and well-being were all similar between night and day shifts (all p > 0.05, **Table 2**).

**Table 2.**
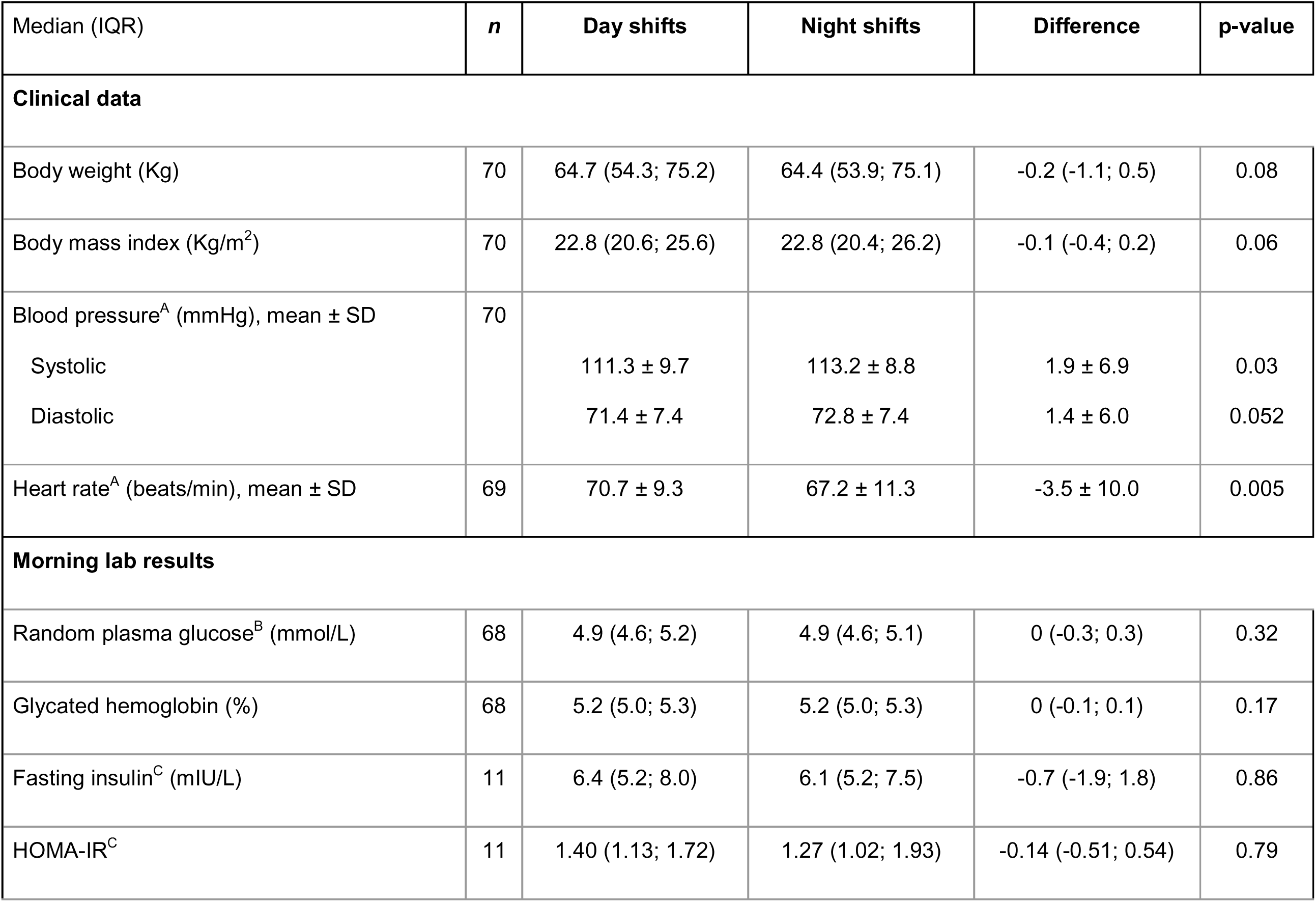
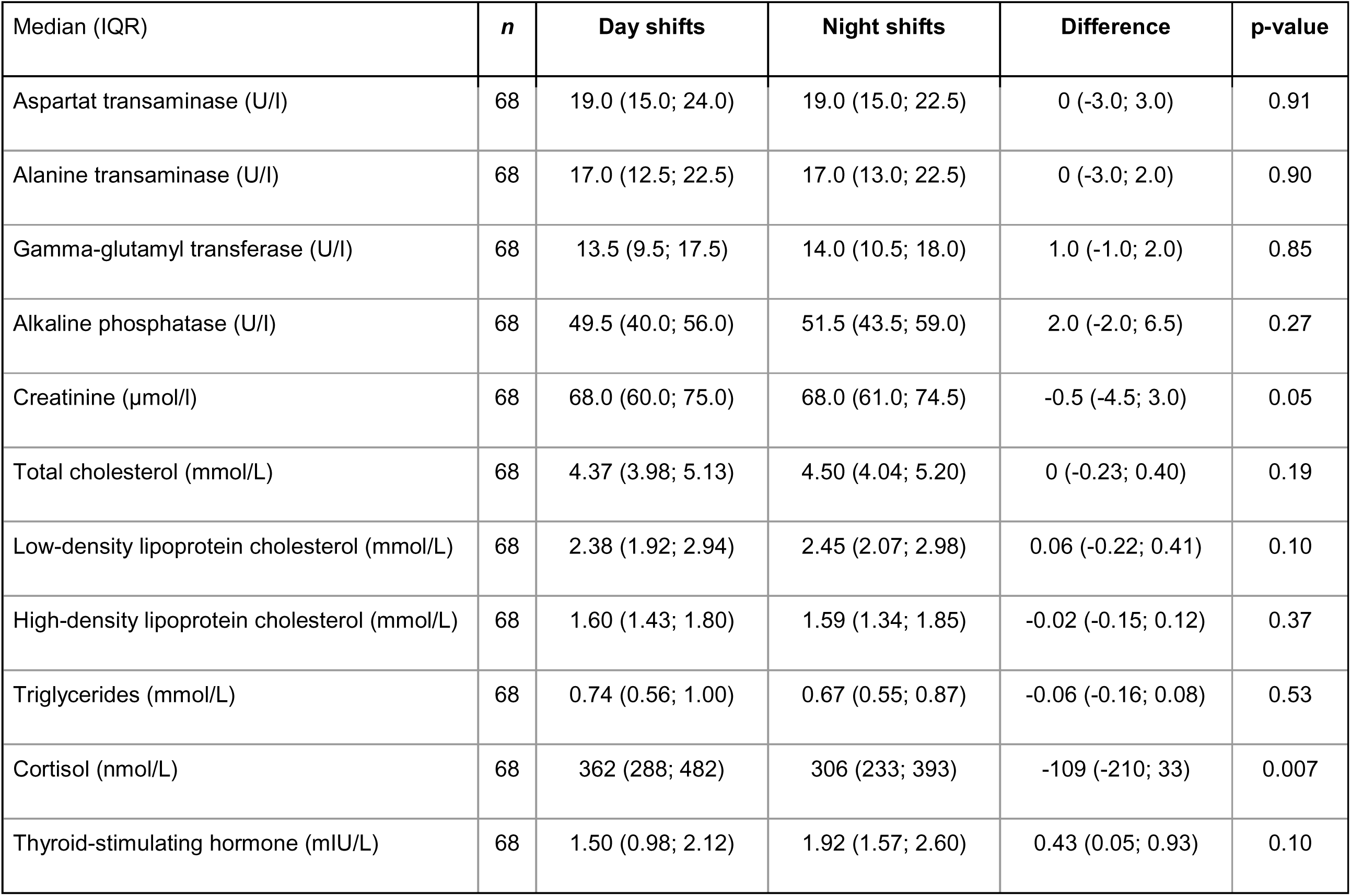
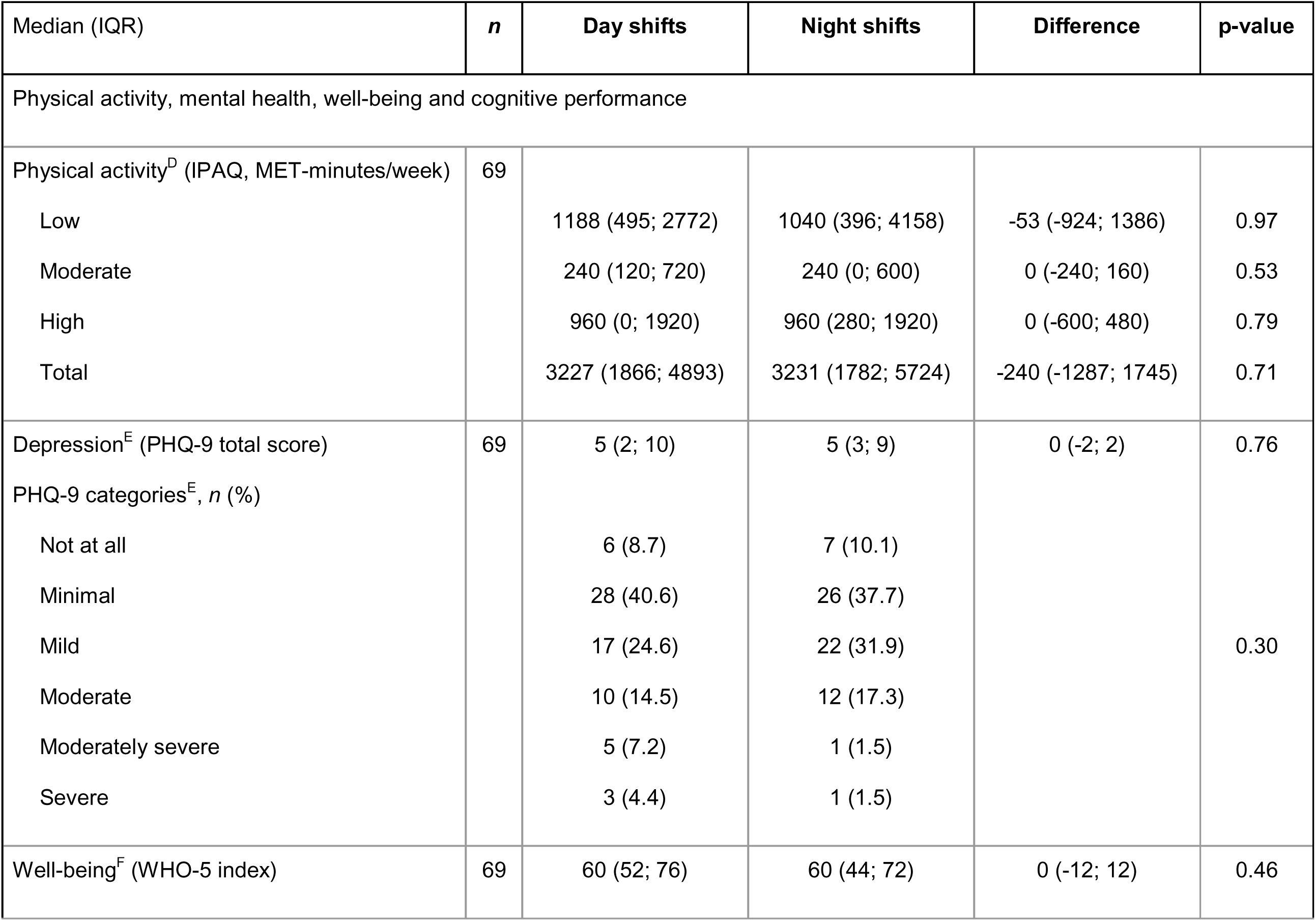
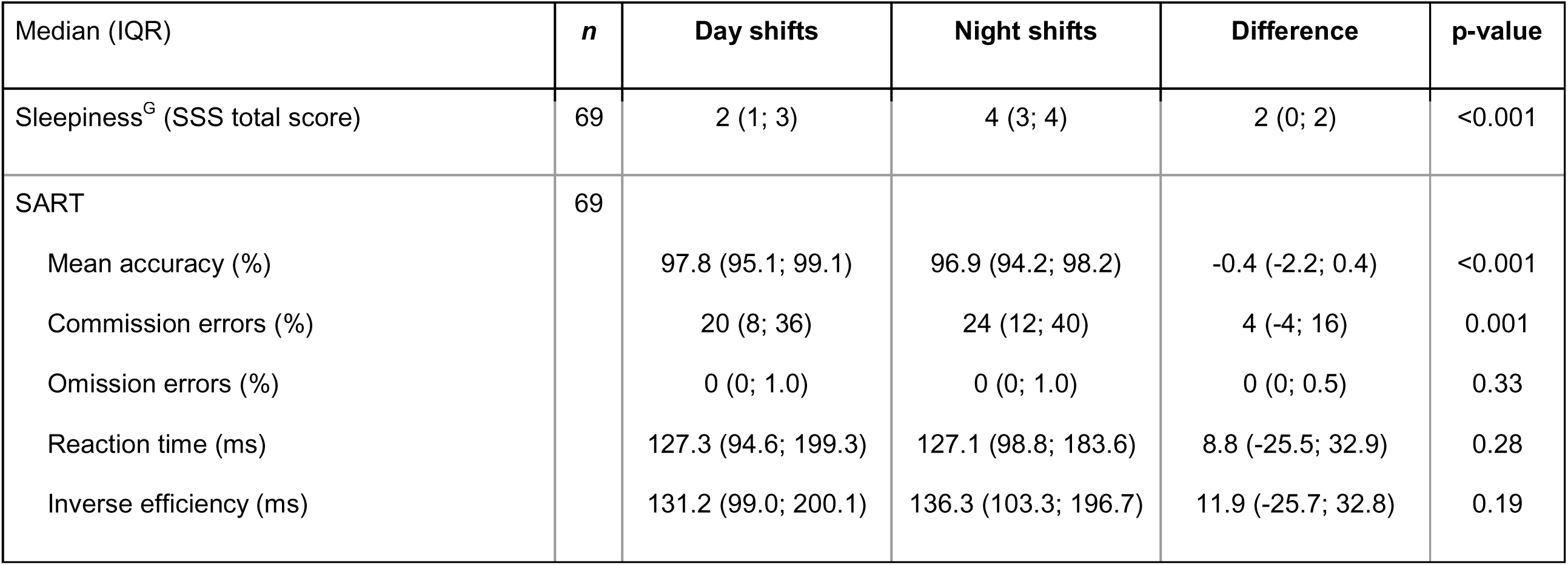
Comparison of cardiometabolic parameters and health outcomes between night and day shifts. ^A^To account for circadian variation, values were compared between readings undertaken at the same time in the day (i.e., in the morning prior to the start of day shift rotation versus in the morning following the end of night shift rotation). ^B^One shift worker had a fasting plasma glucose within the prediabetes range, as defined by the American Diabetes Association (77). ^C^Only those who had fasting measurements after the series of day and night shifts are shown. 66 participants were fasted for day shifts with median fasted insulin of 6.6 (IQR 5.3; 8.6) mIU/L. ^D^Physical activity assessed with the short form of the International Physical Activity Questionnaire (IPAQ) (65), with a total score and three subscores for low, moderate, and high physical activity levels. ^E^Depression assessed with the Patient Health Questionnaire-9 (PHQ-9) (67), score ranging between minimal (1-4 points), mild (5-9 points), moderate (10-14 points), moderately severe (15-19 points), and severe depression (20-27 points). ^F^Well-being assessed with the WHO-5 index (66) ranging from 0 to 100; where 0 represents the worst possible and 100 the best possible quality of life. ^G^Sleepiness assessed with the Stanford Sleepiness Scale (SSS) (68) on a 7-point scale, 1 representing feeling alert and 7 indicating being almost in reverie. See also Supplemental Table 3. <u>Abbreviations</u>: HOMA-IR, Homeostatic Model Assessment for Insulin Resistance; IPAQ, International Physical Activity Questionnaire; MET, Metabolic Equivalent; PHQ-9, Patient Health Questionnaire-9; SSS, Stanford Sleepiness Scale; SART, Sustained Attention to Response Task (10); WHO-5, WHO-5 Well-being Index.

### Night shifts led to poorer cognitive performance and sleep quality with higher levels of fatigue

Cognitive performance was compared between night and day shifts using the Sustained Attention to Response Task (SART) (10) at the end of both shifts (**Table 2**). Night shift work was associated with lower mean accuracy (-0.4%, IQR -2.2 to 0.4, p<0.001) and higher commission errors (4%, IQR -4 to 16, p=0.001) compared to day shifts. However, we found no significant differences in omission errors, reaction time, and inverse efficiency. Higher levels of sleepiness were reported post-night shift compared to post-day shift (+2 on a 7-point scale, IQR 0 to 2, p<0.001) just before performing SART.

Self-reported sleep and fatigue between night and day shifts are compared in **Supplemental Table 3**. Shift work schedules significantly affected the timing of the main sleep-wake pattern (both p<0.001), and participants reported taking more naps during night shifts compared to day shifts (p<0.001). Poorer sleep quality was more commonly reported during night shifts compared to day shifts, as well as insufficient sleep, inadequate restfulness after sleep, and waking earlier than intended (all p<0.001). In our previous analysis of sleep diaries of the same cohort (9), sleep duration (i.e., main sleep period) remained stable across day shifts, whereas during night shifts, sleep duration was significantly longer prior to the first night shift but shorter on subsequent night shifts (all p<0.001). In that previous analysis, we observed similar trends for sleep quality between night and day shifts (9). In terms of fatigue, a higher proportion of participants reported feeling tired during night shifts compared to day shifts (p<0.001).

### Night shifts reduced calorie, protein, and fat intake, with unchanged carbohydrate intake and altered 24-hour timing of consumption

Night shifts were associated with significant temporal changes in food and drink consumption according to the time of day, with higher nighttime calorie intake and reduced lunchtime intake (**Figure 1A**). These trends were overall similar across calories, carbohydrates, protein, fat (**Figure 1A-D**), and fiber (**Supplemental Figure 2**). However, relative nighttime carbohydrate consumption was higher than protein and fat during night shifts as opposed to day shifts. Visualization of the hourly consumption rate comparing carbohydrates and protein (**Figure 1B-C**) showed that relative nighttime carbohydrate intake was higher than protein intake during night shifts. Considering total consumption per day (i.e. the area under the curve of the consumption rates in **Figure 1** upper panels), night shifts were associated with slightly lower consumption of calories (mean -164 ± 530 kcal/day, p=0.015, **Figure 1E**), protein (mean -9.6 ± 28.1 g/day, p=0.007, **Figure 1G**) and fat (mean -8.9 ± 29.3 g/day, p=0.017, **Figure 1H**), and similar carbohydrate intake (mean -3.6 ± 59.2 g/day, p=0.62, **Figure 1F**).

**Figure 1.**
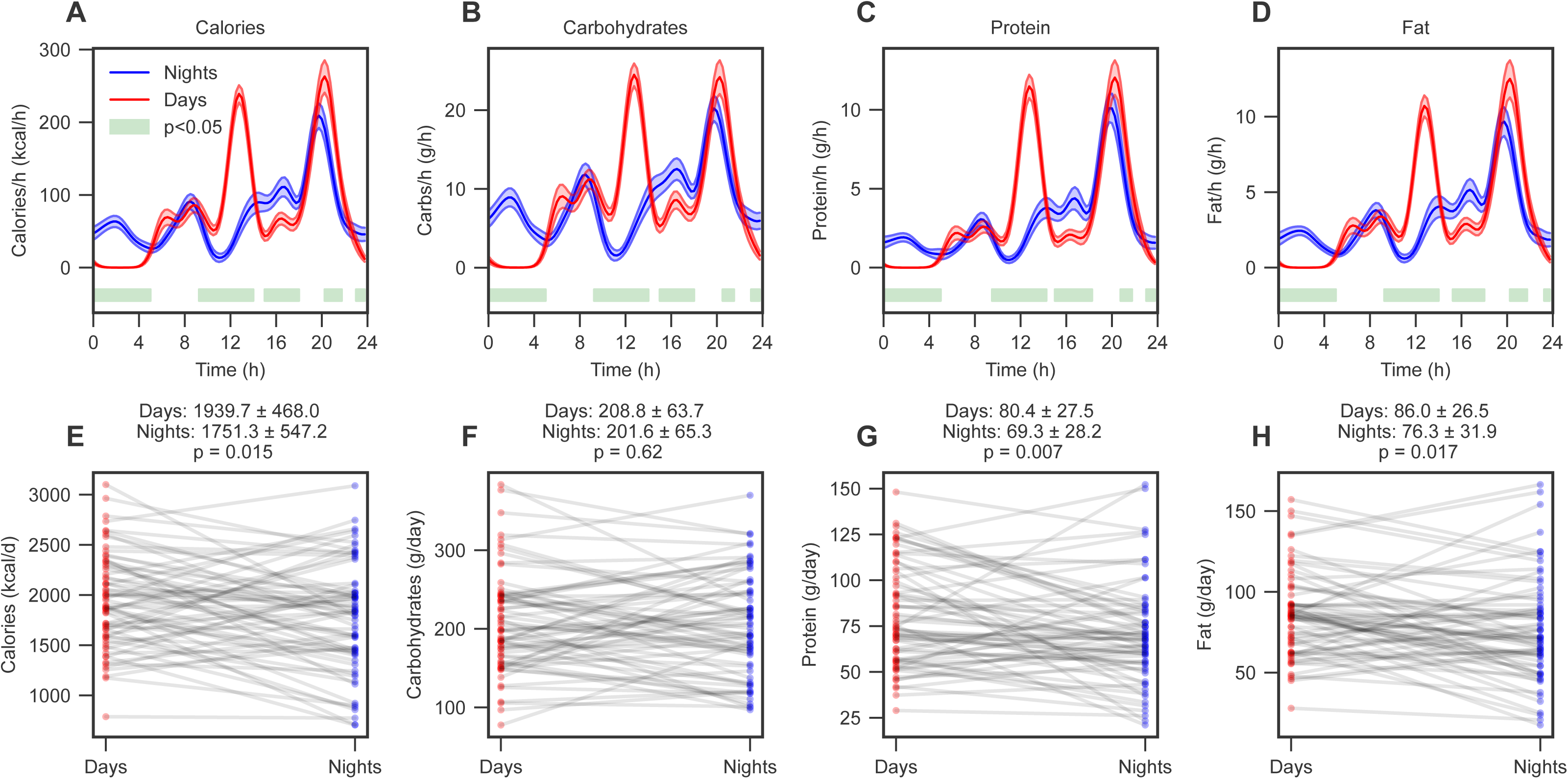
Night shifts were associated with lower calorie, protein, and fat intake, but similar carbohydrate intake compared to day shifts, with significant changes in the timing of consumption across 24 hours. (A-D) The mean consumption rate per hour for night shifts (blue) and day shifts (red) for total calories, carbohydrates, protein, and fat, respectively. The shaded area represents the mean ± SEM, and the green bar represents significant group differences as assessed from p<0.05 in a Wilcoxon non-parametric test, adjusted for multiple testing with the Benjamini-Hochberg procedure. (E-H) Individual average daily intake for the day and night shift period for total calories, carbohydrates, protein, and fat, respectively. Group means ± SD are indicated above each panel. *n* = 66 for all analyses. See also Supplemental Table 4 and Supplemental Figure 2.

This resulted in a higher proportion of carbohydrates during night shifts (+2.6%, IQR -1.3 to 8.6, p=0.002, **Supplemental Table 4**). In comparison to the nutritional reference values for Switzerland (shown in **Supplemental Table 4**) (11), the proportion of total calories from carbohydrate intake was slightly lower during day shifts. In both shift rotations, protein intake met the daily recommendation, whereas the proportion of fat intake exceeded the recommendation. While the intake of dietary fiber was slightly lower during night shift compared to day shift (-1.93 ± 6.36 g/day, p=0.017), both did not meet the daily recommendation of 30 g.

These objective data corroborate the self-reported eating habits between day and night shifts (**Supplemental Table 5**). Lower subjective appetite (p=0.008) and a reduced daily number of meals (p=0.01), along with irregular meal times (p=0.03), were reported during night shifts compared to day shifts. Dinner became the main meal of the day during night shifts (p=0.004), with a higher frequency of eating in front of a screen during night shifts (p=0.001). Lower consumption of alcohol was reported during night shifts compared to day shifts (p<0.001), as confirmed with the smartphone food diary app (**Supplemental Figure 2**).

### Lower 24-hour mean glucose and increased nocturnal glycemic variability during night shifts

The CGM measures glucose as a function of the time of day across multiple study days (example shown in **Figure 2A**), which allows us to estimate how the mean and short-term variation vary according to clock time, using e.g. a moving average. The moving average glucose levels comparing night and day shifts differed significantly as a function of the time of day, with the minimum night shift glucose levels at midday strongly diverging from the maximum glucose levels at 14:00h during day shifts (**Figure 2B**). The moving average glycemic variability differed significantly between shifts across multiple time blocks (green blocks, **Figure 2C**), with a higher nocturnal coefficient of variation (CV) during night shifts but a lower CV in the early afternoon, possibly caused by lunch-skipping behavior during night shifts (**Figure 1A**). Considering all data across the entire shift periods, night shifts were associated with a moderate decrease in the global mean glucose levels (difference -0.19 ± 0.61 mmol/L, p=0.02, **Figure 2D**) and a moderate increase in glycemic variability as assessed with both the global CV (difference +1.45% ± 4.5%, p=0.02, **Figure 2E**) and the mean amplitude of glycemic excursions (MAGE: difference +0.21 ± 0.64 mmol/L, p=0.02, **Figure 2F**) (12).

**Figure 2.**
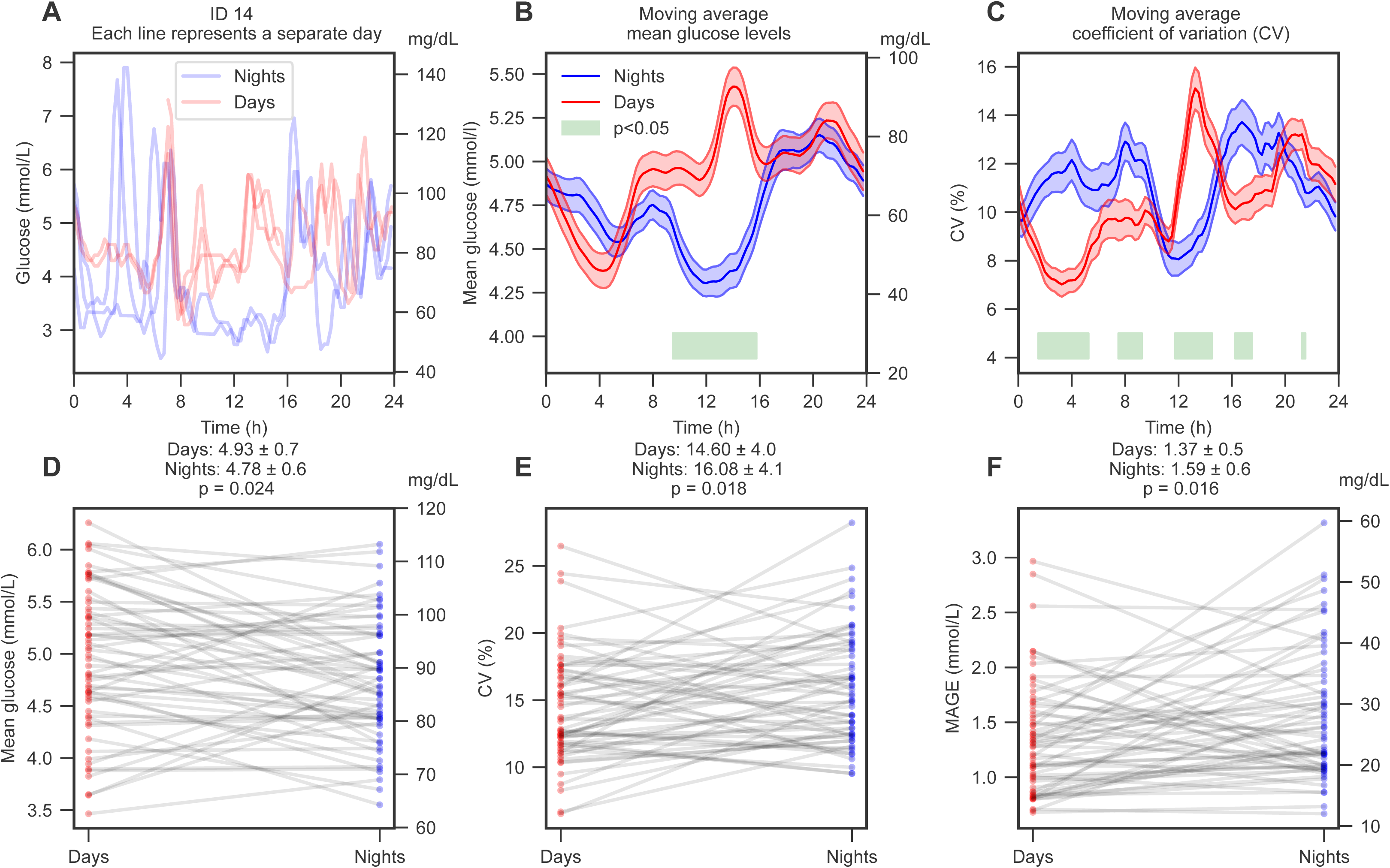
Night shift work is associated with significant differences in the glycemic mean and coefficient of variation (CV) across the time of day, with overall reduced glucose levels but higher glycemic variability. (A) Example of glucose levels as a function of the time of day for one participant (ID 14), where the different lines represent different study days. Blue, night shifts; red, day shifts. (B-C) The 2-hour moving average estimate over the 24-hour clock of glucose mean and CV, respectively. The shaded area represents the mean ± SEM, and the green bar represents significant group differences as assessed from p<0.05 in a Wilcoxon non-parametric test, adjusted for multiple testing with the Benjamini-Hochberg procedure. (D-F) Individual changes for glucose mean, CV, and mean amplitude of glycemic excursions (MAGE) (12), respectively. Group means ± SD are indicated above each panel. *n* = 58 for all analyses. See also Supplemental Table 6.

### Night shift work is associated with lower baseline glucose levels but more pronounced postprandial glucose spikes

We next used a mathematical model of glucose levels (illustrated in **Figure 3A**) that models glucose levels as a superposition of (1) meal-induced glucose spikes (orange lines) using the timestamped food and drink consumption entries recorded with the MyFoodRepo smartphone app (dotted gray lines), and (2) a 24-hour rhythm in the underlying glucose levels (black line) (13). The lowest point (i.e. the trough, nadir) of the baseline glucose levels was lower by -0.29 ± 0.69 mmol/L during night shifts compared to day shifts for the same study participants (p=0.003, **Figure 3B**). Furthermore, the 24-hour baseline glucose levels were significantly shifted during night shifts (p<0.001), with the peak time during afternoon on day shifts (red star, **Figure 3C**) and late evening during night shifts (blue star, **Figure 3C**). The model also revealed that the mean meal height (i.e. the glucose increase following a meal) was 0.23 ± 0.38 mmol/L higher during night compared to day shifts (p<0.001, **Figure 3D**). These differences are illustrated in one example, who exhibits a lower trough in glucose levels but higher average postprandial glucose spikes during night shifts compared to day shifts (**Figure 3F** vs. **Figure 3E**, respectively).

**Figure 3.**
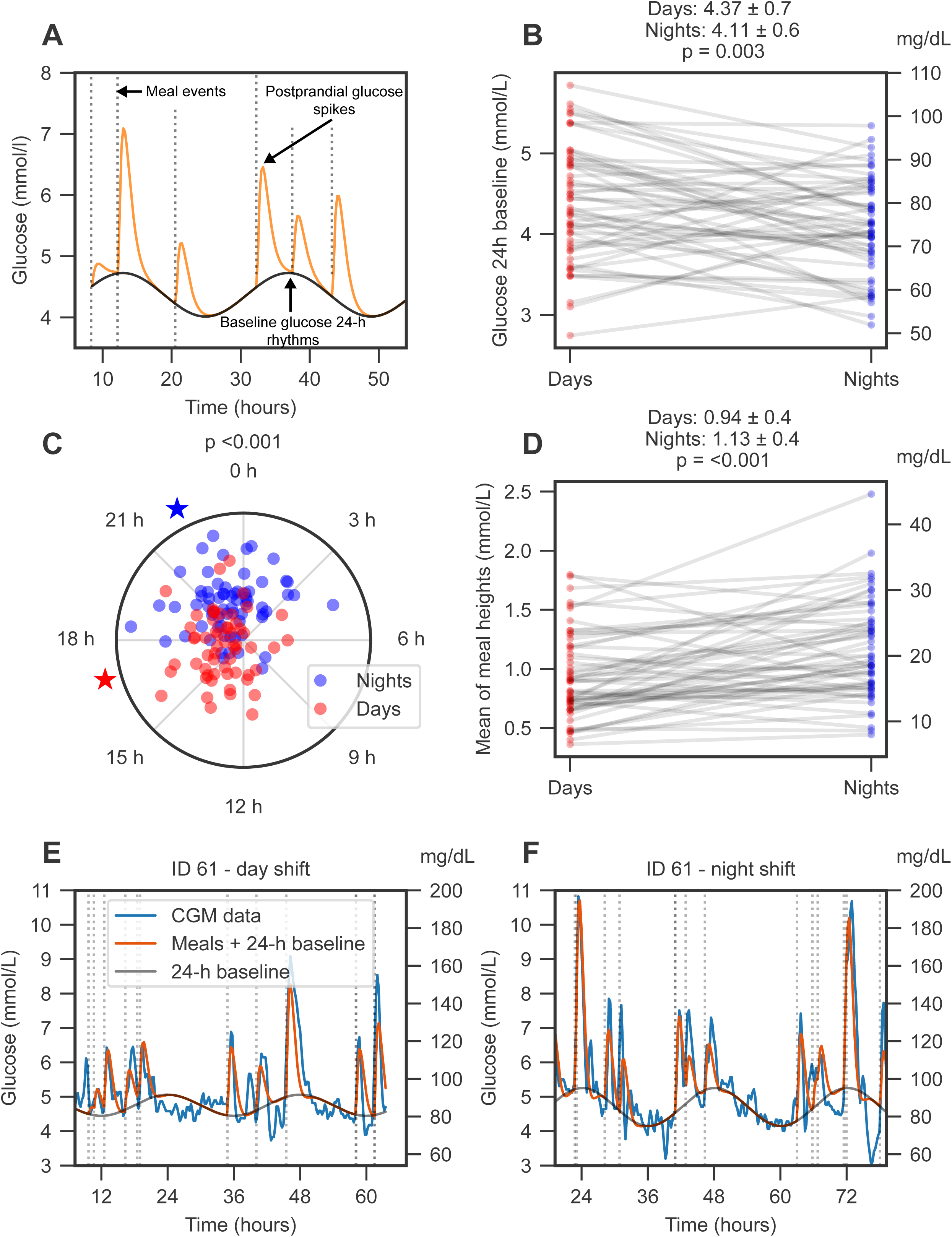
Dynamical modeling of glucose levels reveals that night shift work is associated with a lower baseline glucose level and higher average postprandial glucose spikes. (A) Schematic of the mathematical framework, which uses meal events (dotted gray) to model glucose levels as a superposition of postprandial meal spikes (orange) on top of an underlying 24-hour rhythm (black) (13). (B) Individual changes in the baseline of the glucose 24-hour rhythm. Group means ± SD are indicated above each panel. (C) The peak time of the 24-hour rhythm during night and day shifts. Blue star, mean peak time of night shifts; red star, mean peak time of day shifts; p-value derived from a Kuiper test. (D) Individual changes in the mean glucose response heights after meal consumption. (E-F) An individual example (ID 61) of continuous glucose monitor (CGM) data and the corresponding model fit during day and night shifts. Blue, CGM raw data; orange, model prediction incorporating ingestion events and the underlying 24-hour glucose rhythm; black, the underlying 24-hour glucose rhythm; dashed lines, meal timestamps. *n* = 55 for all analyses. See also Supplemental Table 6.

### Night shifts disrupt 24-hour heart rate and parasympathetic activation rhythms

The activity monitor and electrocardiogram ActiHeart^®^ provided continuous measurements around the 24-hour clock for physical activity, HR, and heart rate variability (HRV) (examples of one day of measurements shown for one participant in **Figure 4A-C**). Continuous actigraphy monitoring showed significantly modified activity profiles across the time of day, with night shifts leading to higher nighttime and lower daytime activity levels compared to day shifts (**Figure 4D**). Activity levels were similar during the early morning (06:00h to 08:00h) and late afternoon/evening (17:00h to 22:00h) periods. The continuous HR data revealed significant disruptions in 24-hour HR fluctuations (**Figure 4E**). The changes to HR showed an overall similar pattern as activity counts (**Figure 4D**) but with one notable difference: while the activity levels were comparable between night and day shifts at 08:00h, the HR was lower during night shifts. This low morning HR in night shifts corroborates the independent morning measurement of resting HR with an arm-cuff BP monitor (see above, **Table 2**). In parallel, we also measured HRV, which reflects parasympathetic (vagal) activity and autonomic nervous system regulation. Daytime HRV was significantly higher around midday on night shifts compared to day shifts (**Figure 4F**), indicating perturbed 24-hour parasympathetic activation.

**Figure 4.**
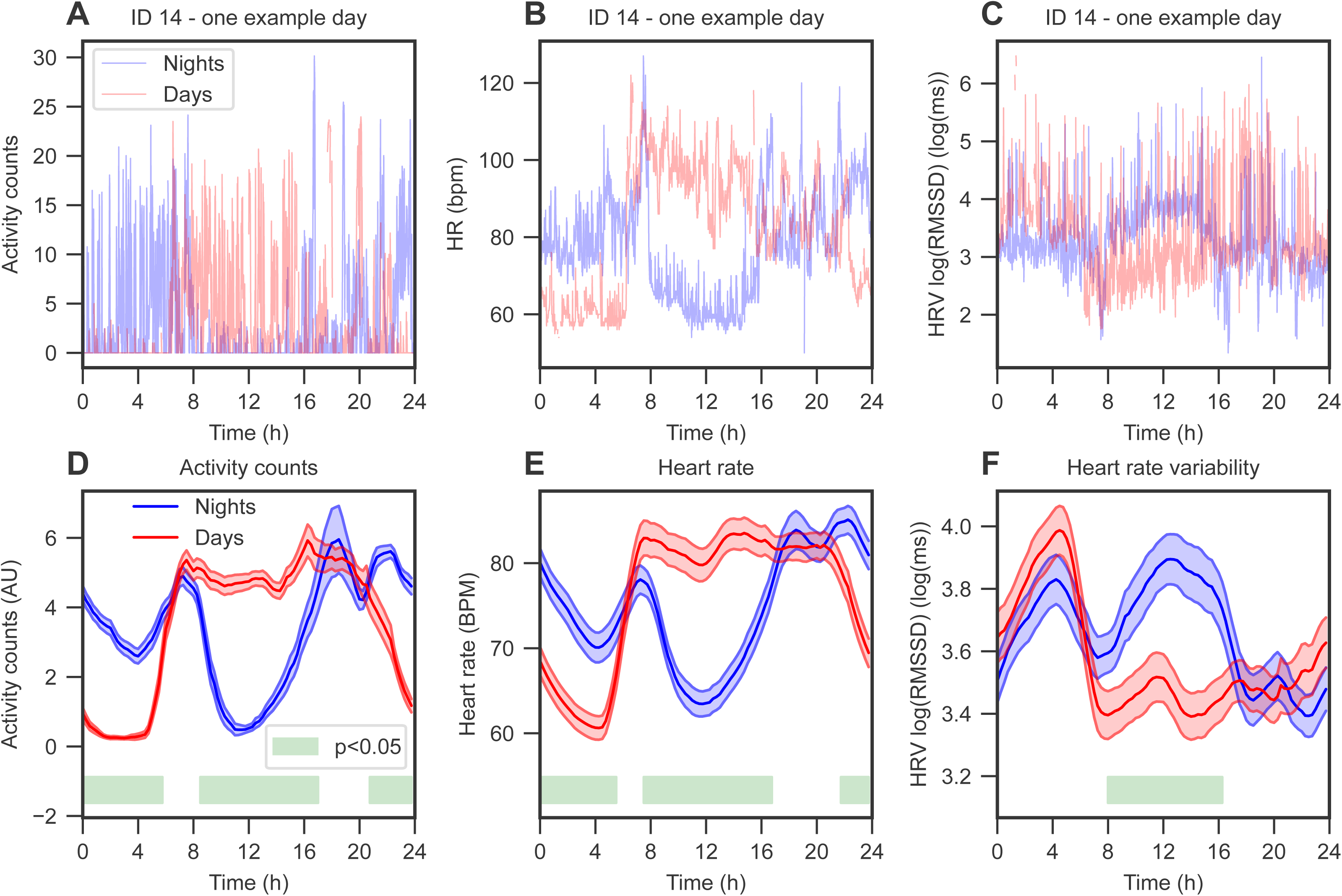
The mean 24-hour profiles of activity (in arbitrary units, AU), heart rate (HR) and heart rate variability (HRV) are perturbed during night shifts. (A-C) Examples from participant ID 14 of one day of measurements for activity counts, HR, and HRV, respectively. Blue, night shifts; red, day shifts. (D-F) The 2-hour moving average over the 24-hour clock of activity counts, HR, and HRV (log(RMSSD)), respectively. Blue, night shifts; red, day shifts. The shaded area represents the mean ± SEM, and the green bar represents significant group differences as assessed from p<0.05 in a Wilcoxon non-parametric test, adjusted for multiple testing with the Benjamini-Hochberg procedure. *n* = 43 for all analyses. See also Supplemental Table 6.

### A higher protein diet and the avoidance of nighttime eating are associated with lower glycemic variability in shift workers

To explore potential nutritional interventions in the shift worker population, we quantified the relationships between the macronutrient composition and nighttime eating with markers of glycemic variability. Among the macronutrients, a higher proportion of carbohydrates was associated with a higher glucose CV (p=0.03, **Figure 5A**). Conversely, a higher protein intake correlated with reduced glucose fluctuations, reflected by a lower CV (p=0.002, **Figure 5B**) and decreased MAGE (p=0.003, **Figure 5F**). Fat intake percentage was not significantly associated with CV (p=0.53, **Figure 5C**) or MAGE (p=0.92, **Figure 5G**). To explore the correlation of nighttime eating with glycemic variability, we defined a continuous parameter as the time of the last calorie intake prior to 05:00h (5 AM) and found that those who ate throughout the night exhibited higher CV (p=0.037, **Figure 5D**) and MAGE (p=0.008, **Figure 5H**). To put these relationships in a clinical context, if these associations were causal then an intervention at 30% protein intake would be predicted to reduce average night-shift MAGE levels from 1.59 to 0.98 mmol/L, and an intervention restricting eating between 20:00h and 05:00h would be expected to reduce night-shift MAGE from 1.59 to 1.33 mmol/L.

**Figure 5.**
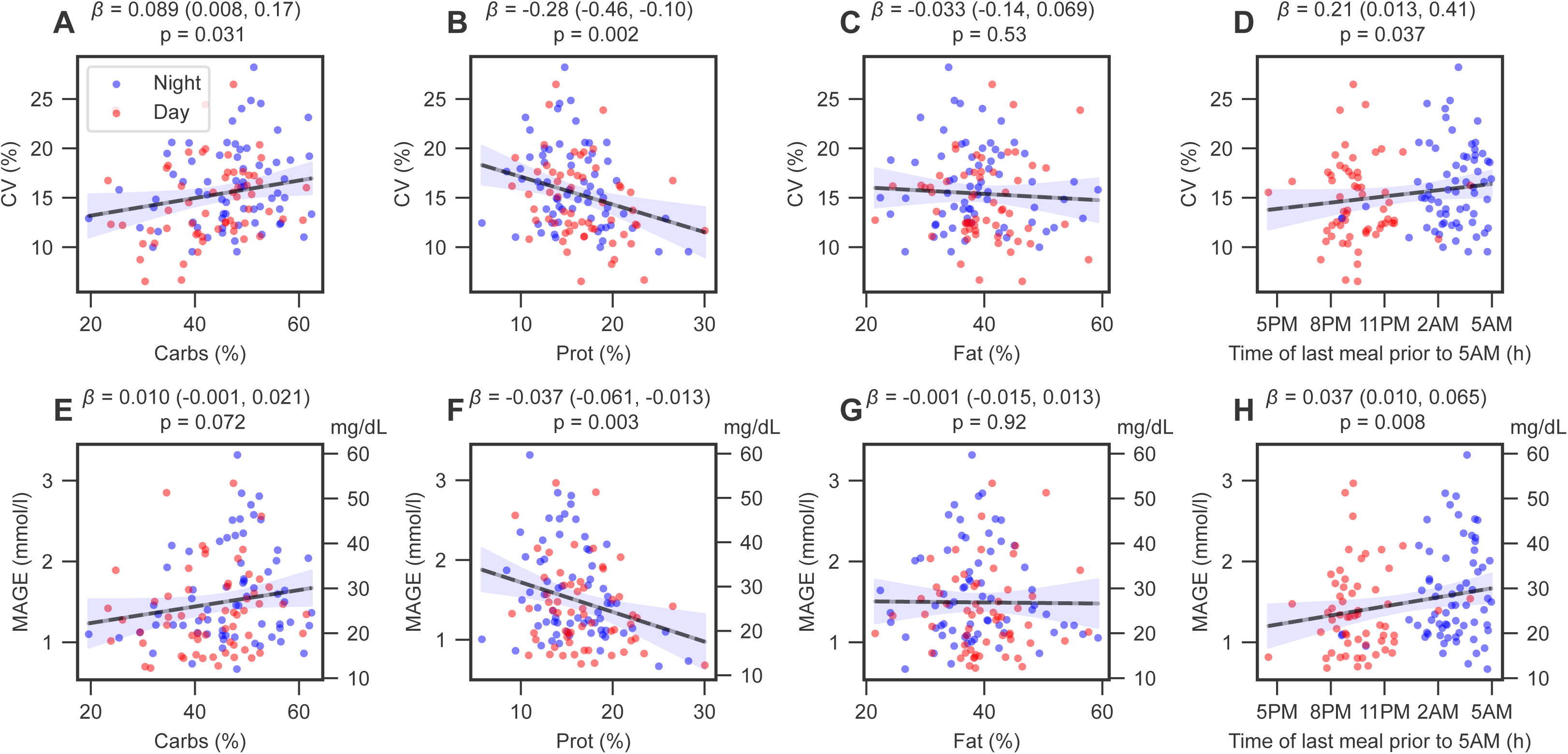
Glycemic variability is associated with increased carbohydrates, lower protein, and nighttime eating. In a linear mixed model pooling day (n=59) and night shifts (n=67), glucose coefficient of variation (CV) and mean amplitude of glycemic excursions (MAGE) (12) were associated with macronutrient proportions (CV: panels A-C; MAGE: panels E-G) and nighttime eating (panels D and H, respectively). Blue dots, night shifts; red dots, day shifts. *β* denotes the inferred regression coefficient (with a 95% confidence interval), which is also shown graphically (gray line and shaded blue, respectively).

## DISCUSSION

In this study of shift workers in their real-world occupational environment, night shift work was associated with several deleterious outcomes. These included disturbances in sleep, increased fatigue, impaired cognitive performance, and adverse effects on cardiometabolic health. A significant change was observed in the dietary behaviors of the same participants between night and day shift periods, particularly in caloric and macronutrient intake, as well as meal timing. These changes may have, in part, contributed to alterations in glucose metabolism during the night shift periods. Our exploratory analyses suggest that the altered glucose regulation could potentially be counteracted by adjusting meal composition and timing during the night shift periods.

Sleep deprivation and disturbances are prevalent among shift workers due to the imposed rhythms that are in desynchrony with their endogenous circadian clocks (3). Expanding on our recent findings from the same cohort of shift workers (9), we now report that night shift work is associated with poor sleep quality, insufficient sleep, inadequate restfulness, and waking earlier than intended. These findings are consistent with the earlier work by Boivin *et al*. (3). Sleep disturbances have implications for job performance, as it leads to fatigue, impaired cognitive function, such as attention and executive performance, and burnout (14, 15). Indeed, we found increased levels of tiredness and decreased levels of attention, as demonstrated by the poorer performance on the SART, post-night shifts compared to post-day shifts. Several pharmacological and non-pharmacological interventions, such as melatonin use and napping strategies, have shown promise in reducing sleep disturbances among shift workers (16–19). In the current cohort, approximately 15.3% of the participants reported using melatonin to aid sleep and more frequent napping during night shifts compared to day shifts.

Altered circadian patterns of blood pressure and autonomic nervous system (higher HR and lower HRV) resulting from repeated exposure to night shifts have been proposed as one of the mechanisms for the increased risk of cardiovascular disease and mortality (20). A meta-analysis of 72 observational studies demonstrated that night shift work is associated with a 1.10 to 1.31-fold increased risk of hypertension (21). Our data reflect this trend, showing higher morning systolic blood pressure in the morning following night shifts compared to the morning following non-night shifts. Blood pressure follows a circadian rhythm, rising upon awakening in the morning and dipping by 10-20% during nighttime hours and sleep (22). However, this nocturnal blood pressure dipping is blunted in night shift workers with a delay in recovery post-night shifts (20, 23–25). Both HR and HRV also exhibit a day-night rhythm. HR increases during the day and decreases at night, whereas HRV shows the opposite pattern, decreasing during the day and increasing at night (26, 27). As shown by our data, these circadian patterns of the autonomic nervous system were perturbed during night shifts. This imbalance in autonomic nervous system regulation could also be explained by the changes in behavior factors during night shift periods, such as sleep, physical activity, and posture (28). In a recent meta-analysis, shift workers exhibited elevated sympathetic nervous system activity, as suggested by reduced HRV (29).

Shift work affects cortisol secretion patterns (30). Our observation of lower morning cortisol levels following night shifts, compared to mornings after non-night shifts, corroborates the findings reported by Ernst *et al*. (31). It has been suggested that the depressed cortisol level post-night shifts may be due to a reversal in cortisol rhythm, with the timing of morning cortisol rise shifted to a later time in a day (31, 32). Approximately two days off work are required to restore the altered circadian rhythm of cortisol secretion (33), however, we did not measure cortisol during time off after night shifts in our study for logistic reasons.

A recent meta-analysis concluded that there are no intraindividual differences in 24-hour energy intake between day shifts and night shifts (34). Of the seven included studies in a within-participant comparison, two found higher energy intake on night shifts (35, 36) and two others reported no difference (37, 38). Three studies reported lower intake on night shifts (39–41), which are consistent with our findings. The mean difference in energy intake between shifts observed in our study (−163.8 kcal/day) is comparable to that reported by Bouillon-Minois *et al.* (−206 kcal/day) (41). Work-related stress-induced food restriction has been suggested as a contributing factor to the reduced energy intake observed during night shifts (41). Our findings support this notion, as shift workers in the present study reported lower subjective appetite scores and fewer meals consumed during night shifts compared to day shifts. Our macronutrient analysis showed that the lower energy intake during night shifts was due to reduced consumption of protein and fat, but not carbohydrates. This is in line with previous studies showing trends towards low protein (36, 39–41) and fat intake (40, 41) during night compared to day shifts. The availability and accessibility of high energy-dense snacks during night shifts explained the majority of energy intake that comes from carbohydrate consumption (42). Previous studies showed lower intake of fruit and vegetables among night shift workers than day workers (43, 44), which is consistent with our findings of lower dietary fiber intake during night shifts compared to day shifts, but both were below the recommended 30 g of daily intake. The intake of fruit and vegetables should be encouraged in shift workers given the important role of dietary fiber for the prevention of cardiometabolic disease (45).

While total energy and macronutrient intake in shift workers have been extensively studied (34), the temporal distribution of calorie and macronutrient intake in relation to shift schedules remains largely underexplored (46). In the present study, meal times shifted significantly during night shifts, with more calories and macronutrients consumed at night compared to day shifts. Indeed, following a night shift, shift workers spent the daytime sleeping (9), which often led to skipping lunch. Our findings on self-reported eating habits supported this, as the majority of participants indicated that dinner became the main meal during night shifts, whereas lunch was the main meal during their day shifts. Additionally, participants reported that skipping meals, irregular eating patterns, and increased snacking were the most common dietary adjustments in response to the night shift schedule, consistent with previous qualitative research among shift workers (42).

Both laboratory-controlled and epidemiological studies have demonstrated that night shift work (circadian misalignment) leads to increased fasting plasma glucose, impaired glucose tolerance, higher postprandial glucose concentrations, elevated insulin levels, and decreased insulin sensitivity (4, 5, 7, 47, 48). Our study extends these findings by showing that night shift work contributes to increased glycemic variability with higher postprandial glucose spikes, relative to day shifts. This is important, as even in the absence of diabetes, higher glycemic variability is a risk factor for coronary atherosclerosis and may predict incident cardiovascular events and type 2 diabetes (49, 50). The biological mechanisms underlying shift work-induced impaired glucose regulation remain poorly understood, but several potential mechanisms have been proposed. Morris *et al.* demonstrated that in healthy adults undergoing a circadian misalignment protocol, lower glucose tolerance in the evening was mediated by reduced β-cell function and decreased insulin sensitivity (5). In a later study involving circadian-misaligned adults, Wefers *et al.* found that decreased muscle insulin sensitivity resulted from reduced skeletal muscle non-oxidative glucose disposal. Furthermore, misalignment of the muscle molecular clock favors intramuscular fatty acid metabolism over glucose metabolism, leading to disrupted energy metabolism (7). In another study, Sharma *et al.* found that the higher postprandial glucose concentrations observed during night shifts were due to impaired β-cell responsiveness to glucose (48). Taken together, these findings may explain the higher glycemic variability and postprandial glucose spikes observed during night shift periods in our study.

In this study, we found that not only the timing of meals but also the composition of meals was associated with glycemic variability. In particular, higher daily carbohydrate intake correlates with higher glycemic variability, whereas higher daily protein intake correlates with lower glycemic variability. Consuming proteins alongside carbohydrate-rich meals during the day can effectively lower postprandial blood glucose peaks (51, 52). Furthermore, consuming a high protein and low carbohydrate meal at night significantly reduced postprandial glucose excursions by 71.4% (53). This effect is driven by enhanced insulin sensitivity, influenced by the insulinotropic properties of dietary proteins and slower gastric emptying (54, 55). Regarding meal timing, we found that nighttime eating (defined as consumption up until 5 AM) was associated with high glycemic variability. Our study expands upon a previous observational study in shift workers, which found that eating at night (21:00h to 06:00h) led to significantly higher insulin levels compared to consuming most calories during the daytime (06:00h to 21:00h) (56). Additionally, studies indicate that blood glucose levels after a meal are significantly higher when food is consumed at night compared to the same meal eaten in the morning (57–59).

The main strength of OPTI-SHIFT is its real-world setting, comparing the same participants’ 24-hour behavioral and physiological patterns between the night and day shift periods, complementing findings from epidemiological and laboratory-controlled studies. The within-participant comparison also eliminates many confounders associated with interindividual variability in between-participant comparisons. Additionally, the homogeneity of the study population and setting (i.e. 96% of healthcare workers) reduced confounders associated with the heterogeneity of shift work patterns across various occupational settings. Also, the use of wearable devices for continuous measurements provided in-depth longitudinal high-resolution insights into 24-hour changes in cardiometabolic parameters compared to a single time-point assessment. The use of the MyFoodRepo food diary app to assess temporal eating patterns throughout shift schedules that include timing of meals has significantly enhanced the accuracy of data collection, compared to routinely used self-reporting food diaries (35–41).

The study population consists of generally healthy young shift workers with normal BMI, despite our efforts to recruit participants with a diverse range of comorbidities and BMI categories. This limits the generalizability to other shift worker populations with different health statuses, professions, and occupational settings. Despite the homogeneity of the present cohort (i.e. 96% of healthcare workers), there remains a wide variability in job responsibility (physical and mental demands), years of exposure to night shift work, and shift work patterns (most often irregular vs. regular and permanent night shifts). All these factors have been shown to influence individual adaptation to circadian misalignment (3). The focus of this study was on the acute, short-term physiological effects of shift work, and other study designs would be required to investigate its long-term cardiometabolic effects. Within our study, the total duration of shift work was highly correlated (Pearson coefficient ρ=0.85) with age, a known cardiometabolic risk factor, and once age was adjusted for, we did not see a significant association between long-term shift work duration and any of the components of metabolic syndrome in our young and healthy population. The present study did not measure blood pressure and cortisol levels continuously throughout the day to limit the burden on participants, which would have provided a more comprehensive understanding of their variations and fluctuations over time. In addition, the majority of participants did not fast before the post-night shift blood tests; therefore, some lab results should be interpreted with this consideration in mind. Data collection did not extend to after the series of night shifts, which could have provided more insights into the lasting effects of circadian misalignment on the measured parameters. The interval between the two shift periods ranged from weeks to several months for organizational reasons, thus occurring in different seasons, and might have biased the comparison between shifts: for day shift rotation, 47 were in summer and 23 in winter, and for night shift rotation, 32 were in summer and 40 in winter (9).

In conclusion, night shift work is associated with adverse cardiometabolic health and cognitive outcomes. There is an urgent need to implement lifestyle countermeasures to improve the health and well-being of shift workers. Our findings suggest that modifying macronutrient intake and meal timing during night shift work may be beneficial. Therefore, further investigation through randomized controlled trials assessing these dietary strategies to improve the cardiometabolic health of night shift workers is now warranted.

## METHODS

### Sex as a biological variable

Sex was not considered as a biological variable.

### Study design and participants

OPTI-SHIFT was a prospective, observational, single center study undertaken at Geneva University Hospitals (HUG), Switzerland. Participants were recruited via posters, announcements, oral presentations, social media, and the hospital website. Data collection was conducted from January 2021 to August 2024.

Participants were assessed at two time points to compare the effects of a series of three consecutive night shifts (circadian misalignment) to a series of three consecutive day shifts (circadian alignment, **Supplemental Figure 1**). Each time point consisted of two visits, and participants thus attended four study visits over the whole study. At each time point, the set-up visits (visits 1 and 3) occurred at least 24 hours before a series of night or day shifts to ensure that measurements were accurate to cover the series of shifts, i.e. the accuracy of continuous glucose monitors (CGM) tends to improve 12-24 hours after insertion. The follow-up visits (visits 2 and 4) took place after a series of night shifts and after a series of day shifts, respectively. The second time point was scheduled at least two weeks after the last night shift or change in time zone, to ensure that circadian clocks were resynchronized (i.e. end of circadian misalignment). The interval between the first and second time points was kept ideally under three months, for feasibility and to reduce the effects of daylight and seasons on measurements.

#### Inclusion criteria

- Clinical criteria

- ○ Men and women
- ○ Age of 20-50 years (to avoid too much variability in the circadian clock parameters)
- Work-related criteria

- Working ≥ 80% full-time equivalent over the previous month and during the study
- Working night shifts (i.e. shifts that comprise working hours between 23:00h and 06:00h) in rotation with day shifts (i.e. shifts that comprise working hours between 06:00h and 20:00h)
- Planned to work at least three consecutive night shifts during the study preceded by at least a day off or day shift
- Planned to work at least three consecutive day shifts during the study
- Confident use of a smartphone compatible with the study application (iOS, Android) and able to regularly take pictures of consumed food/drinks
- Able to give fully informed consent and follow the study procedures

#### Exclusion criteria

- Clinical criteria

- Planned or current pregnancy during the study
- Menopausal women (due to changes in sleeping habits, hormonal status, and sometimes the impact on weight and mental concentration)
- Major illness or hospitalization over the previous month
- Carries a pacemaker, pump, or other medical devices that can be disabled by a magnet
- Major mental illness
- Study-related criteria

- Trip to a different time zone (≥ 2-hour time difference) over the previous month or planned during the study
- Enrolled in an interventional clinical trial (potentially interfering with the main outcomes) over the previous month or planned during the study

### Outcomes

*Questionnaires.* We recorded socio-demographic data (e.g. age, sex, ethnicity), and work-related variables (e.g. work pattern, shift work system, motivations, workload) using a subset of questions adapted from the Standard Shiftwork Index (SSI) (60). We used the REDCap platform for data entry and questionnaires (61, 62). We assessed the self-reported sleep duration, sleeping times, and sleep quality using the Pittsburgh Sleep Quality Index (PSQI) (63). The PSQI has seven components (subjective sleep quality, sleep latency, sleep duration, sleep efficiency, sleep disturbances, use of sleeping medications, and daytime dysfunction), with each component score ranging from 0 to 3, with the total score ranging from 0 to 21, where a higher score indicates poorer sleep quality. The chronotype was assessed using the Morningness-Eveningness Questionnaire (MEQ), with scores ranging from ‘definitely morning’ (70–86 points), ‘moderately morning’ (59–69 points), ‘neutral’ (42–58 points), ‘moderately evening’ (31–41 points), to ‘definitely evening’ (16–30 points) (64). Physical activity levels were assessed using the short form of the International Physical Activity Questionnaire, with a total score and three subscores for low, moderate, and high physical activity levels (65). Well-being was evaluated with the World Health Organization-Five Well-being Index with scores ranging from 0 to 100; where 0 represents the worst possible, whereas 100 represents the best possible quality of life (66). Mental health was evaluated by the Patient Health Questionnaire-9, with scores ranging from minimal (1-4 points), mild (5-9 points), moderate (10-14 points), moderately severe (15-19 points), and severe (20-27 points) depression (67). The Stanford Sleepiness Scale (SSS) evaluates the level of sleepiness at a specific time in the day that uses a 7-point scale, 1 representing feeling alert and 7 indicating being almost in reverie (68). Self-reported eating habits were collected using a questionnaire and subjective appetite was measured using a visual analog scale ranging from 0 (not hungry) at all to 10 (very hungry).

*Sustained Attention to Response Test (SART).* Cognitive performance was assessed using the Sustained Attention to Response Task (SART) (10) on the online platform PsyToolkit (69, 70), right after the SSS questionnaire (68) (see above, **Supplemental Figure 1**). This is a 5-minute standard test undertaken when participants are seated in front of a computer screen in a quiet room. Participants were presented with an integer between 1 and 9, appearing 225 times in random order with different sizes. Participants have to press a key when they see a digit on screen but withhold it when they are presented with the number ‘3’, thus testing reaction time and inhibition. Outcomes are reported as mean accuracy, commission errors (a key was pressed when no key should be pressed, i.e. after a ‘3’, the ‘NO-GO trials’), omission errors (lack of key press when it should have been pressed, i.e. after a digit other than a ‘3’, the ‘GO trial’), reaction time (ranging between 0 and 1500 milliseconds; where lower values indicate better performance), and inverse efficiency (a combined measure of speed and accuracy with a higher score indicates poorer performance).

*Body composition and vital signs.* Weight was measured in light clothing, and the body mass index (BMI) was calculated as the weight in kilograms divided by the height in meters squared (**Supplemental Figure 1**). Fat mass and fat-free mass were estimated from the electric resistance and reactance measured by BIA using the Nutriguard-M^®^ device (Data Input, Germany). Skin adhesive electrodes were placed on the right hand and foot, while the subject was lying on their back. An alternating electrical current of 50 kHz and 0.8 mA was applied. The measured resistance and reactance were used to estimate fat-free mass by a formula developed in Geneva and validated against dual energy X-ray absorptiometry (71). Fat mass was calculated by subtracting fat-free mass to weight. Fat mass index and fat-free mass index were calculated by dividing them by the height squared. Waist and hip circumferences and waist-to-hip ratio (WHR) were assessed according to the WHO standard procedure (72). Blood pressure was measured three times with a calibrated monitor (Omron Intellisense BP monitor, Omron Healthcare, USA) and an appropriately sized arm cuff, after 5 min of rest in the sitting position, and the last two values were averaged.

*Blood measurements.* Fasting plasma glucose, glycated hemoglobin (HbA1c), lipid profile (total, HDL and LDL cholesterol, triglycerides), liver function tests, cortisol, and full blood count were measured with the routine assays at the clinical lab of HUG. Blood draw at the end of the night shifts could also be performed in the non-fasting state to reduce the burden to study participants.

*MyFoodRepo smartphone application.* The MyFoodRepo smartphone food diary app (Digital Epidemiology Lab, EPFL) recorded the timestamps of consumed food and drink along with pictures, barcodes of packaged items, or free text descriptions (73, 74). The macronutrient composition of each consumed item was automatically extracted from a constantly updated nutritional composition table, verified by trained dieticians.

*Continuous Glucose Monitors (CGM).* CGM data was recorded with an Abbott FreeStyle Libre^®^ Pro device (Abbott Diabetes), which reports interstitial glucose every 15 mins. During the initial period of recording, the CGM data can occasionally spend the majority of the time at the device minimal value of 2.2 mmol/L due to the transient effects of microtrauma, local pressure leading to lower blood flow and device calibration, and in this case, we removed the initial data until the device exited this floor state. As a preliminary step of glucose dynamics modeling (see below), abnormally low glucose values (that are typical of the early recording hours) were also corrected by removing long timescale trends within the time series. CGM metrics related to glycemic control were extracted from the CGM data using the R package rGV (75), and we focused on the mean glucose level, coefficient of variation (CV = SD/mean), and the mean amplitude of glycemic excursions (MAGE) (12).

*Chest-worn activity and heart monitor.* The CamNTech ActiHeart^®^ device (version 5, CamNTech, United Kingdom) assessed physical activity (activity counts, proprietary algorithm), heart rate (HR, beats/min), and heart rate variability (HRV, using log root mean square of successive differences [log(RMSSD)] between normal heartbeats) (76).

### Statistics

*Calculating 24-hour physiological profiles using a 2-hour rolling average.* To calculate the moving average 24-hour profile for glucose, activity counts, HR, and HRV, we first defined grid points spanning from 0 to 24 hours every 15 minutes. We then computed the average values over a 2-hour window centered on each grid point. For glucose, the common definition of coefficient of variation (CV = SD/mean) is calculated over all data points. We thus used this definition to calculate the 2-hour moving average CV by dividing the 2-hour moving average standard deviation by the 2-hour moving average mean at each grid point. For the nutritional variables, the mean consumption rate per hour was calculated by using a kernel density estimate (with a von Mises distribution, *κ* = 30) and then normalizing such that the total area under the curve equaled the daily consumption rate.

*Modeling ingestion events and glucose data.* We combined the ingestion events with CGM data based on a recently published computational framework (13). The food and drink consumption entries recorded with the MyFoodRepo smartphone app are used to model postprandial interstitial glucose increases, and both the height of the glucose spikes (termed “response heights”) and the time needed for glucose to return to baseline (termed “response half-life”) are inferred for each individual and each shift condition independently. The model also includes an underlying 24-hour cosinor function to describe 24-hour oscillations in baseline (i.e. irrespective of meal intake) glucose levels, which is a mathematical model that assumes baseline glucose levels follow a sinusoidal pattern over the course of a day. This 24-hour cosinor function is described by the three parameters ‘baseline’, ‘peak-to-trough amplitude’, and ‘peak time’, which are similarly learned for each participant and each shift condition independently. The model was trained by minimizing the error between the model and the CGM data using a maximum *a posteriori* probability (MAP) estimate from 200 different starting conditions, and the settings and parameters of the original method were left unchanged.

*Missing data handling.* The MyFoodRepo smartphone app data were considered missing when fewer than 700 calories per day were recorded. The threshold of 700 calories was chosen empirically based on manual inspection of the MyFoodRepo and CGM data showing multiple glucose spikes without corresponding meal entries below this threshold. The CGM metrics were calculated when at least 48 hours of data were recorded. We calculated metrics from the first 48 hours of the MyFoodRepo and CGM data to ensure a comparable measurement window between day and night shifts and to avoid oversampling of certain times of the day in either group. The MyFoodRepo data were analyzed alongside the CGM data when both MyFoodRepo data and CGM data were concomitant and not missing. Upon data extraction, the ActiHeart^®^ produces a quality estimation of the signal (range 0-1), and we filtered the data based on the threshold of 0.8. The ActiHeart^®^ data was used when at least 2000 valid 1-minute time points were recorded, and the 2-hour rolling average was considered missing when fewer than 10 valid measurements were available for each 2-hour window.

*Statistical analyses.* Continuous and normally distributed variables are reported as mean ± SD and compared with paired t-tests using the matched day and night data for each participant. In this within-participant comparison, we thus did not adjust for variables such as age, sex, body composition, and physical activity. Normality was assessed with the Shapiro-Wilk test. Non-normally distributed continuous variables are reported as median (IQR) and compared with the Mann–Whitney U test (i.e. Wilcoxon rank-sum test). Categorical variables are presented as the number of participants (% in group) and compared using the chi-squared test. Associations between the nutritional and glycemic variables were quantified by combining the day and night shift data within a linear mixed regression model accounting for repeated data in individual participants, where the glycemic variables were considered as the dependent variables and the nutritional variables (calories, macronutrient proportions, and nighttime eating) were considered as the independent variables. The aim of the random effect was to account for potential correlations within the same participant, and the model was fitted using the ‘MixedLM’ class from the ‘statsmodels’ Python library (v0.13.0). We performed a complete-case analysis using all available data and hence no imputation methods were used for missing data. As this was an exploratory observational study, the sample size was not predetermined and multiple testing corrections were not performed, unless stated otherwise. Nevertheless, we aimed to include a large sample of individuals to maintain statistical power, allowing for a sufficient margin to account for invalid data due to compliance and technological issues commonly encountered in observational studies involving wearable devices and mHealth apps.

### Study approval

The study protocol was approved by the Ethics Committee of the Canton of Geneva (approval no. 2021-01008) and is registered on ClinicalTrials.gov (no. NCT05177965). This study was carried out in accordance with Good Clinical Practices and the Swiss Human Research Act. All participants provided written informed consent prior to enrollment in the study.

## Data availability

Further information and requests for resources, data, and code should be directed to the lead contact and principal investigator, Tinh-Hai Collet.

## Data Availability

All data produced in the present study are available upon reasonable request to the corresponding author.

## AUTHOR CONTRIBUTIONS

The order of the co-first authors was determined on the basis of the significance, time, and effort that each author invested in the project. Conceptualization, C.P., C.D. and T.H.C.; Methodology, N.E.P., A.H., C.P., C.D. and T.H.C.; Software, N.E.P., V.S., M.S. and T.H.C.; Formal analysis, F.C.J., N.E.P. and T.H.C.; Investigation – data collection, A.H., C.J. and T.H.C.; Data curation, F.C.J., N.E.P., A.H., C.J. and T.H.C.; Visualization, N.E.P. and T.H.C.; Writing – original draft, F.C.J. and N.E.P.; Writing – review and editing, all authors; Funding acquisition, C.P., C.D. and T.H.C.; Resources and Supervision, C.P. and T.H.C.

## FUNDING SUPPORT

This project was supported by the Nutrition 2000*plus* Foundation and the Medical Board of the Geneva University Hospitals. F.C.J. is supported by a grant from the Swiss Government Excellence Scholarship (2024.0465). N.E.P. is supported by grants from the Swiss Society of Endocrinology and Diabetes, the Hjelt Foundation, the Panacée Foundation, and the Swiss Multiple Sclerosis Society. A.H. and C.J. are supported by grants from the EPFL. A.D.B. is supported by the SwissLife *Jubiläumsstiftung* Foundation. S.L.H.’s and M.A.’s research is supported by the European Union’s Horizon 2020 research and innovation programmed under the Marie Sklodowska-Curie Innovative Training Networks (ITN, grant no. 860613). C.D.’s research is supported by grants from the Swiss National Science Foundation (310030–219187), the Leenaards Foundation, the Vontobel Foundation, the Novartis Foundation for Medical-Biological Research, Gerthruda-Meissner Foundation, the SwissLife *Jubiläumsstiftung* Foundation, Velux Foundation, Swiss Cancer League, ISREC Foundation, Academic Society of Geneva (SACAD), Ernst and Lucie Schmidheiny Foundations, and Ligue Pulmonaire Genevoise. T.H.C.’s research is supported by grants from the Swiss National Science Foundation (32003B-212559), the Leenaards Foundation, the Vontobel Foundation, the SwissLife *Jubiläumsstiftung* Foundation, and the Swiss Society of Endocrinology and Diabetes.

## ACKNOWLEDGMENTS

The authors wish to thank all participants, the clinical and research team at the Clinical Research Center, Geneva University Hospitals and Faculty of Medicine, University of Geneva, the laboratory team at the HUG Sérothèque (Biobank), the team at Digital Epidemiology laboratory, EPFL, and the annotators of all recorded collected food and drink pictures via the MyFoodRepo app. The authors have not used any large language models to generate the text, tables, and figures of this manuscript.

## Notes

**Conflict of interest statement** The authors have declared that no potential conflict of interest exists.

**Funding.** This project was supported by the Nutrition 2000plus Foundation and the Medical Board of the Geneva University Hospitals.

### Competing Interest Statement

The authors have declared no competing interest.

### Clinical Trial

NCT05177965

### Funding Statement

This project was supported by the Nutrition 2000plus Foundation and the Medical Board of the Geneva University Hospitals. F.C.J. is supported by a grant from the Swiss Government Excellence Scholarship (2024.0465). N.E.P. is supported by grants from the Swiss Society of Endocrinology and Diabetes, the Hjelt Foundation, the Panacée Foundation, and the Swiss Multiple Sclerosis Society. A.H. and C.J. are supported by grants from the EPFL. A.D.B. is supported by the SwissLife Jubiläumsstiftung Foundation. S.L.H's and M.A.'s research is supported by the European Union's Horizon 2020 research and innovation programmed under the Marie Sklodowska-Curie Innovative Training Networks (ITN, grant no. 860613). C.D.'s research is supported by grants from the Swiss National Science Foundation (310030-219187), the Leenaards Foundation, the Vontobel Foundation, the Novartis Foundation for Medical-Biological Research, Gerthruda-Meissner Foundation, the SwissLife Jubiläumsstiftung Foundation, Velux Foundation, Swiss Cancer League, ISREC Foundation, Academic Society of Geneva (SACAD), Ernst and Lucie Schmidheiny Foundations, and Ligue Pulmonaire Genevoise. T.H.C.'s research is supported by grants from the Swiss National Science Foundation (32003B-212559), the Leenaards Foundation, the Vontobel Foundation, the SwissLife Jubiläumsstiftung Foundation, and the Swiss Society of Endocrinology and Diabetes.

### Author Declarations

The Ethics Committee of the Canton of Geneva gave ethical approval for this work (BASEC registry no. 2021-01008).

### Summary of Updates

Abstract and section formatting; figure S2 updated

