## Supplemental Material for "The nutritional and metabolic impact of night shift work in a real-world setting"

**Supplemental Table 1.** Participant characteristics (*related to Table 1*).

| **Participant characteristics** | ***n* = 72** |
|---|---|
| Educational level, *n* (%) | |
| College, High school | 3 (4.2) |
| Apprenticeship | 5 (6.9) |
| Professional school | 39 (54.1) |
| University, University of applied science | 21 (29.2) |
| Other | 4 (5.6) |
| Household composition, median (IQR) | |
| Number of children | 0 (0; 1) |
| Age of youngest child, years | 3.5 (1.5; 7.0) |
| Age of oldest child, years | 6.0 (3.6; 12.3) |
| Number of older incumbent persons | 0 (0; 0) |
| Self-reported ethnicity, *n* (%) | |
| African | 5 (6.9) |
| European | 65 (90.3) |
| North American | 2 (2.8) |
| South American | 3 (4.2) |
| Other ethnicity | 2 (2.8) |
| **Clinical data** | |
| Body mass index categories, *n* (%) | |
| Underweight | 2 (2.7) |
| Normal | 49 (68.1) |
| Overweight | 16 (22.2) |
| Obesity | 5 (7.0) |
| Self-reported comorbid conditions, *n* (%) | |
| Hypertension | 1 (1.4) |
| Dyslipidemia | 3 (4.2) |
| Obstructive sleep apnea | 1 (1.4) |
| Thyroid disorder | 3 (4.2) |
| Asthma | 9 (12.5) |
| Cancer | 2 (2.8) |
| Depression | 3 (4.2) |
| Smoking status, *n* (%) | |
| Current | 11 (15.3) |
| Past | 11 (15.3) |
| Never | 50 (69.4%) |
| Chronotype[A] (*n* = 69), *n* (%) | |
| Definitely morning type | 0 |
| Moderately morning type | 21 (30.4) |
| Neutral type | 43 (62.3) |
| Moderately evening type | 5 (7.3) |
| Definitely evening type | 0 |

Footnotes: [A]Assessed with the Morningness-Eveningness Questionnaire, with a total score ranging between 'definitely morning' (70–86 points), 'moderately morning' (59–69 points), 'neutral' (42–58 points), 'moderately evening' (31–41 points), and 'definitely evening' types (16–30 points) (1).

**Supplemental Table 2.** Work patterns, shift work systems, motivations, and workload.

| **Shift work schedule** | ***n* = 72** |
|---|---|
| **Employment** | |
| Work time, hours | |
| Weekly work duration excluding overtime | 40.0 (35.5; 43.3) |
| Weekly overtime | 1 (0; 2) |
| Commuting time, minutes | |
| Day shift to work | 20 (15; 30) |
| Day shift from work | 20 (15; 30) |
| Night shift to work | 15 (10; 30) |
| Night shift from work | 20 (15; 30) |
| **Shift work system** | |
| No. successive day shifts, days | 3.0 (2.5; 4.0) |
| No. successive night shifts, days | 3.0 (2.5; 3.5) |
| No. successive number of days off | 2.0 (2.0; 2.5) |
| Maximum no. successive shifts of any kind between days off in the past month | 5 (4; 5) |
| No. weekends off per month | 2 (2; 2) |
| No. night shifts in a year, days | 55 (36; 72) |
| Types of night shifts (*n* = 70), *n* (%) | |
| Permanent night shift | 1 (1.4) |
| A single block of night duty per year | 1 (1.4) |
| Occasional blocks of night duty per year | 9 (12.7) |
| A block of nights per month | 11 (15.5) |
| Two blocks of nights per month | 23 (32.4) |
| One or two nights each week | 5 (7.0) |
| Other | 21 (29.6) |
| **Impact of shift work on socio-familial life** | |
| Main reason for working shifts, *n* (%) | |
| Part of the job | 70 (97.2) |
| The only job available | 2 (2.8) |
| More convenient for domestic responsibility | 4 (5.6) |
| Higher rates of pay | 4 (5.6) |
| Others | 8 (11.1) |
| Willingness to give up working shifts over day time job without shifts, *n* (%) | |
| Definitely not | 6 (8.3) |
| Probably not | 12 (16.7) |
| Maybe not | 7 (9.7) |
| Probably yes | 25 (34.7) |
| Definitely yes | 22 (30.6) |
| Partner's weekly paid employment (*n* = 47), hours | 42 (40, 42) |

| **Shift work schedule** | ***n* = 72** |
|---|---|
| Partner's usual work pattern, *n* (%) | |
| Daytime - no shifts | 30 (63.8) |
| Rotating shifts with nights | 10 (21.3) |
| Rotating shifts without nights | 2 (4.3) |
| Permanent night shifts | 2 (4.3) |
| Other | 3 (6.4) |
| Partner's perception of participants' work shifts, *n* (%) | |
| Extremely unsupportive | 2 (4.3) |
| Fairly unsupportive | 3 (6.4) |
| Quite indifferent | 6 (12.8) |
| Fairly supportive | 16 (34.0) |
| Extremely supportive | 20 (42.6) |
| **Self-rated workload during night shifts compared to day shifts** | |
| Physical workload, *n* (%) | |
| Much lighter | 9 (12.5) |
| A bit lighter | 18 (25.0) |
| About the same | 28 (38.9) |
| A bit heavier | 8 (11.1) |
| Much heavier | 9 (12.5) |
| Mental workload, *n* (%) | |
| Much lighter | 5 (6.9) |
| A bit lighter | 20 (27.8) |
| About the same | 24 (33.3) |
| A bit heavier | 13 (18.1) |
| Much heavier | 10 (13.9) |
| Time pressures, *n* (%) | |
| Much lighter | 20 (27.8) |
| A bit lighter | 23 (31.9) |
| About the same | 13 (18.1) |
| A bit heavier | 11 (15.3) |
| Much heavier | 5 (6.9) |
| Emotional stress, *n* (%) | |
| Much lighter | 6 (8.3) |
| A bit lighter | 15 (20.8) |
| About the same | 19 (26.4) |
| A bit heavier | 23 (31.9) |
| Much heavier | 9 (12.5) |

Footnotes: Questions adapted from a subset of questions in the Standard Shiftwork Index (2). Data are presented as median (IQR) for continuous variables, or *n* (%) for categorical parameters.

**Supplemental Table 3.** Comparison of sleep and fatigue between night and day shifts (*related to Table 2*).

| **Sleep habits, sleep disturbances and fatigue (*n* = 72)** | **Day shifts** | **Night shifts** | **p-value** |
|---|---|---|---|
| Preferred sleep duration daily irrespective of shift work, hours | 7.59 (1.07) | | |
| Time of sleep onset | 22:53h (22:00h; 23:00h) | 09:00h (08:30h; 09:23h) | <0.001 |
| Time of wake-up | 06:00h (05:30h; 06:30h) | 14:30h (13:30h; 16:00h) | <0.001 |
| No. naps taken during shifts | 0 (0; 0) | 1 (0; 1) | <0.001 |
| Self-reported sleep quality, *n* (%) | | | <0.001 |
| Extremely badly | 0 | 4 (5.6) | |
| Quite badly | 2 (2.8) | 16 (22.2) | |
| Moderately bad | 20 (27.8) | 29 (40.3) | |
| Quite well | 35 (48.6) | 20 (27.80 | |
| Extremely well | 15 (20.8) | 3 (4.2) | |
| Perception of sleep sufficiency, *n* (%) | | | 0.001 |
| Nowhere near enough | 7 (9.7) | 14 (19.4) | |
| Could do with a lot more | 9 (12.5) | 26 (36.1) | |
| Could do with a bit more | 41 (56.9) | 21 (29.2) | |
| Get the right amount | 14 (19.4) | 11 (15.3) | |
| Get plenty | 1 (1.4) | 0 | |
| Perceived restfulness after sleep, *n* (%) | | | <0.001 |
| Definitely no rested | 4 (5.6) | 12 (16.7) | |
| Not very rested | 16 (22.2) | 33 (45.8) | |
| Moderately rested | 29 (40.3) | 19 (26.4) | |
| Quite rested | 20 (27.8) | 8 (11.1) | |
| Extremely rested | 3 (4.2) | 0 | |
| Difficulty falling asleep, *n* (%) | | | 0.59 |
| Never | 24 (33.3) | 21 (29.2) | |
| Rarely | 16 (22.2) | 26 (36.1) | |
| Sometimes | 23 (31.9) | 16 (22.2) | |
| Frequently | 8 (11.1) | 8 (11.1) | |

| Sleep habits, sleep disturbances and fatigue (*n* = 72) | Day shifts | Night shifts | p-value |
|---|---|---|---|
| Always | 1 (1.4) | 1 (1.4) | |
| Waking earlier than intended, *n* (%) | | | <0.001 |
| Never | 28 (38.9) | 7 (9.7) | |
| Rarely | 21 (29.2) | 7 (9.7) | |
| Sometimes | 16 (22.2) | 19 (26.4) | |
| Frequently | 6 (8.3) | 24 (33.3) | |
| Always | 1 (1.4) | 15 (20.8) | |
| Use of sleeping pills, *n* (%) | | | 0.46 |
| Never | 68 (94.4) | 66 (91.7) | |
| Rarely | 2 (2.8) | 1 (1.4) | |
| Sometimes | 2 (2.8) | 5 (6.9) | |
| Frequently | 0 | 0 | |
| Always | 0 | 0 | |
| Melatonin usage to aid sleep, *n* (%) | | | 0.66 |
| Never | 61 (84.7) | 63 (87.5) | |
| Rarely | 2 (2.8) | 4 (5.6) | |
| Sometimes | 4 (5.6) | 5 (6.9) | |
| Frequently | 3 (4.2) | 0 | |
| Always | 2 (2.8) | 0 | |
| Tiredness during shift, *n* (%) | | | <0.001 |
| Never | 0 | 0 | |
| Rarely | 10 (13.9) | 0 | |
| Sometimes | 32 (44.4) | 15 (20.8) | |
| Frequently | 26 (36.1) | 30 (41.7) | |
| Always | 4 (5.6) | 27 (37.5) | |

Footnotes: Questions adapted from a subset of questions in the Standard Shiftwork Index (SSI) (2). Data are presented as median (IQR) for continuous variables, or *n* (%) for categorical parameters.

**Supplemental Table 4.** Comparison of calorie and macronutrient intake between night and day shifts (*related to Figure 1*).

| Daily consumption (*n* = 66) | Reference values[A] | Day shifts | Night shifts | Difference | p-value |
|---|---|---|---|---|---|
| Calories (kcal/day) | 1861 to 2147 for women<br>2305 to 2672 for men | 1939.7 ± 468.0 | 1751.3 ± 547.2 | -163.8 ± 529.9 | 0.015 |
| Carbohydrates (g/day) | n/a | 208.8 ± 63.7 | 201.6 ± 65.3 | -3.6 ± 59.2 | 0.62 |
| Protein (g/day)<br>Relative to body weight (g/kg) | n/a<br>0.83 g/kg body weight | 74.1 (56.9; 100.9)<br>1.26 (0.86; 1.58) | 67.4 (53.0; 83.1)<br>1.05 (0.80; 1.36) | -9.6 ± 28.1<br>-0.15 ± 0.44 | 0.007<br>0.007 |
| Fat (g/day) | n/a | 86.0 ± 26.5 | 76.4 ± 31.9 | -8.9 ± 29.3 | 0.017 |
| Alcohol (g/day) | 10 g for women<br>20 g for men | 0.0 (0.0; 7.7) | 0.0 (0.0; 0.0) | 0.0 (-6.3; 0.0) | <0.001 |
| Fiber (g/day) | 30 | 19.1 (15.6; 23.9) | 17.8 (13.9; 21.1) | -1.93 ± 6.36 | 0.017 |
| Carbohydrates (%) | 45 to 60 | 43.3 ± 8.6 | 47.0 ± 9.0 | 2.6 (-1.3; 8.6) | <0.001 |
| Protein (%) | n/a | 16.6 ± 4.0 | 15.8 ± 4.1 | -0.6 ± 4.0 | 0.23 |
| Fat (%) | 20 to 35 | 39.7 ± 6.5 | 38.4 ± 7.5 | -1.6 ± 7.5 | 0.09 |
| Alcohol (%) | n/a | 0.0 (0.0; 2.7) | 0.0 (0.0; 0.0) | 0.0 (-2.4; 0.0) | <0.001 |

Footnotes: Data are presented as mean ± SD or median (IQR), depending on the normal distribution of data. [A]Nutritional reference values for healthy adults aged between 18 and 65 in Switzerland (3).

**Supplemental Table 5.** Self-reported eating habits, alcohol and caffeine intake, and tobacco use between night and day shifts.

| **Eating habits**, median (IQR) | ***n*** | **Day shifts** | **Night shifts** | **Difference** | **p-value** |
|---|---|---|---|---|---|
| Self-rated subjective appetite, mean ± SD | 69 | 6.1 ± 2.0 | 5.5 ± 2.1 | -0.7 ± 2.1 | 0.008 |
| Changes in eating habits during night shifts, *n* (%) | 72 | | | | |
| Longer eating duration | | | 10 (13.9) | | |
| Irregular meals | | | 34 (47.2) | | |
| Skipping meals | | | 37 (51.4) | | |
| Unhealthy food consumption | | | 22 (30.6) | | |
| Snacking | | | 32 (44.4) | | |
| Number of meals in the past 24 hours | 70 | 3 (2; 3) | 2 (2; 3) | 0 (-1; 0) | 0.01 |
| Main meal of the day, *n* (%) | 69 | | | | |
| Breakfast | | 6 (8.7) | 3 (4.3) | | |
| Lunch | | 31 (44.9) | 7 (10.1) | | 0.004 |
| Dinner | | 30 (43.5) | 56 (81.2) | | |
| Other | | 2 (2.9) | 3 (4.4) | | |
| Frequency of eating outside the usual mealtime in the past 24 hours | 70 | 1 (0.5; 2) | 2 (1; 3) | 0 (-0.3; 1) | 0.03 |
| Frequency of eating in front of a screen in the past 24 hours | 69 | 1 (0.2; 1) | 1 (1; 2) | 0 (0; 1.5) | 0.001 |
| Alcohol intake (units) | 69 | 0 (0; 2) | 0 (0; 0) | 0 (-1; 0) | <0.001 |
| Caffeine intake (cups) | 56 | | | | |
| Coffee | | 6 (2.75; 8) | 5 (2; 8) | 0 (-1; 1) | 0.62 |
| Tea | | 0 (0; 3) | 1 (0; 2) | 0 (-0.5; 1) | 0.26 |
| Caffeinated soda/ shake drinks | | 0 (0; 0) | 0 (0; 0) | 0 (0; 0) | 0.39 |
| Tobacco use throughout the shift period (numbers) | | | | | |
| Cigarettes | 11 | 15 (4; 40) | 6 (2; 30) | 0 (-9; 4) | 0.79 |
| E-cigarettes | 7 | 0 (0; 0) | 0 (0; 0) | 0 (0; 0) | 0.91 |
| Vaporizers | 8 | 0 (0; 0) | 0 (0; 0) | 0 (0; 0) | 0.93 |

Footnotes: Data are presented as mean ± SD or median (IQR), depending on the normal distribution of data.

**Supplemental Table 6.** Comparison of CGM metrics and Actiheart® measurements between night and day shifts (*related to Figures 2-3-4*).

| | *n* | Day shifts | Night shifts | Difference | p-value |
|---|---|---|---|---|---|
| **CGM metrics** | | | | | |
| Mean glucose (mmol/L) | 58 | 4.93 ± 0.67 | 4.78 ± 0.59 | -0.19 ± 0.61 | 0.024 |
| Coefficient of variation (%) | 58 | 13.85 (11.83; 17.25) | 15.47 (12.72; 18.96) | 1.45 ± 4.52 | 0.018 |
| Mean amplitude of glycemic variation (mmol/L) | 58 | 1.31 (0.99; 1.65) | 1.45 (1.12; 1.99) | 0.21 ± 0.64 | 0.016 |
| Baseline of baseline glucose cosinor model (mmol/L) | 55 | 4.37 ± 0.70 | 4.11 ± 0.59 | -0.29 ± 0.69 | 0.003 |
| Amplitude of baseline glucose cosinor model (mmol/L) | 55 | 0.53 (0.41; 0.68) | 0.62 (0.46; 0.82) | 0.08 ± 0.36 | 0.11 |
| Damping coefficient | 55 | -0.60 (-1.02; -0.29) | -0.74 (-1.32; -0.29) | -0.31 (-0.73; 0.35) | 0.10 |
| Half-life (h) | 55 | 1.25 (1.01; 1.55) | 1.23 (1.04; 1.51) | -0.06 (-0.20; 0.25) | 0.88 |
| Mean of meal heights (mmol/L) | 55 | 0.90 (0.70; 1.16) | 1.06 (0.86; 1.37) | 0.20 (0.01; 0.42) | <0.001 |
| **Actiheart® measurements** | | | | | |
| Activity count | 43 | 3.43 (2.94; 4.12) | 3.11 (2.72; 4.00) | -0.21 (-0.66; 0.24) | 0.12 |
| Heart rate (beats/min) | 43 | 77.54 (72.01; 82.99) | 75.66 (67.39; 79.18) | -1.77 ± 4.63 | 0.018 |
| Heart rate variability [log(RMSSD)] | 43 | 3.58 ± 0.45 | 3.67 ± 0.42 | 0.07 (-0.10 to 0.23) | 0.073 |

Footnotes: Data are presented as mean ± SD or median (IQR), depending on the data normal distribution.

Abbreviations: CGM, Continuous Glucose Monitor; RMSSD, Root Mean Square of Successive Differences.

**Supplemental Figure 1.**

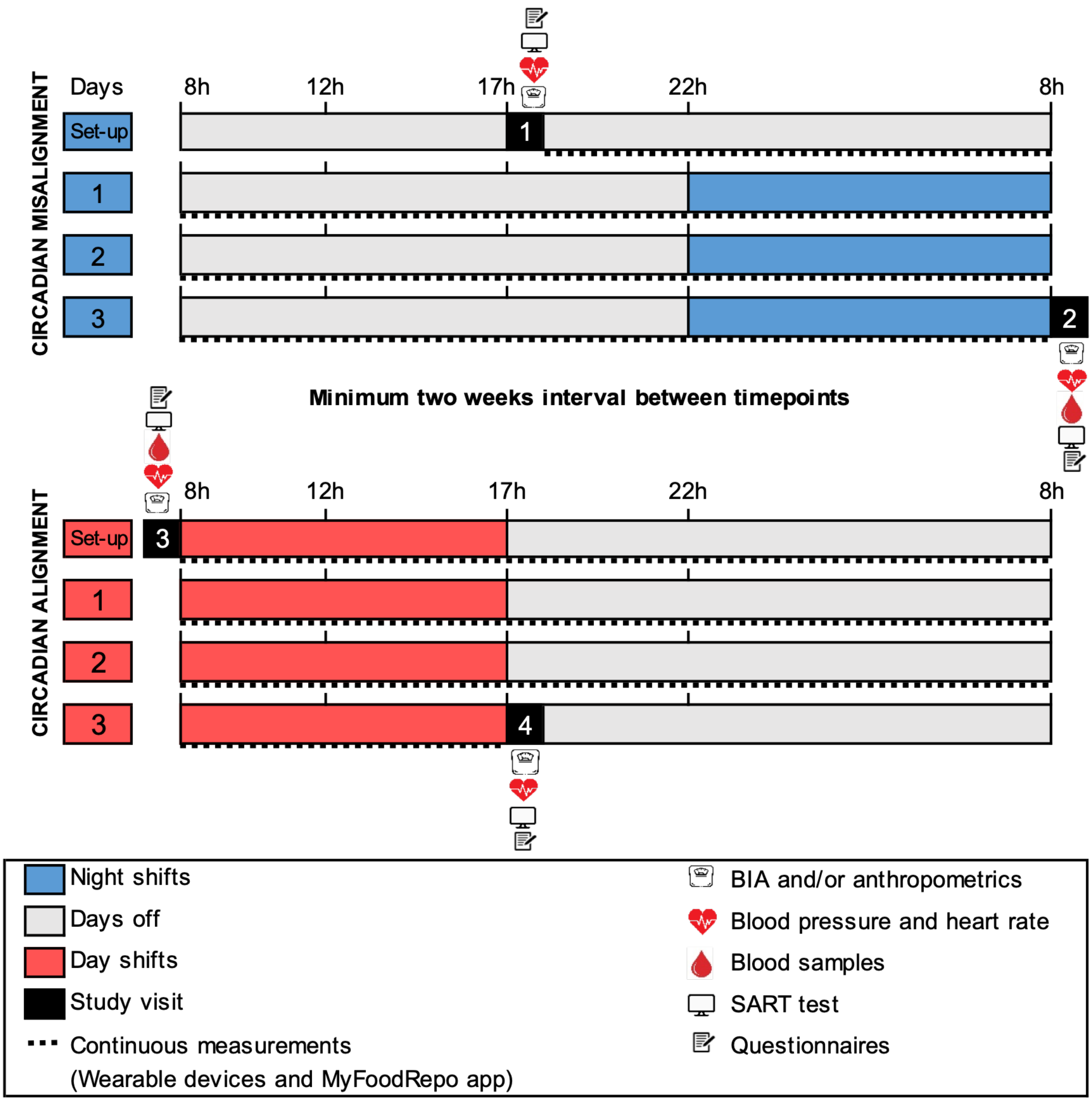


**Figure S1.** Study design of the OPTI-SHIFT observational study. The actual clock times of the shift start and end times could vary slightly according to the individual shift rotations. Abbreviations: BIA, Bioelectrical Impedance; SART, Sustained Attention to Response Task (4).

**Supplemental Figure 2.**

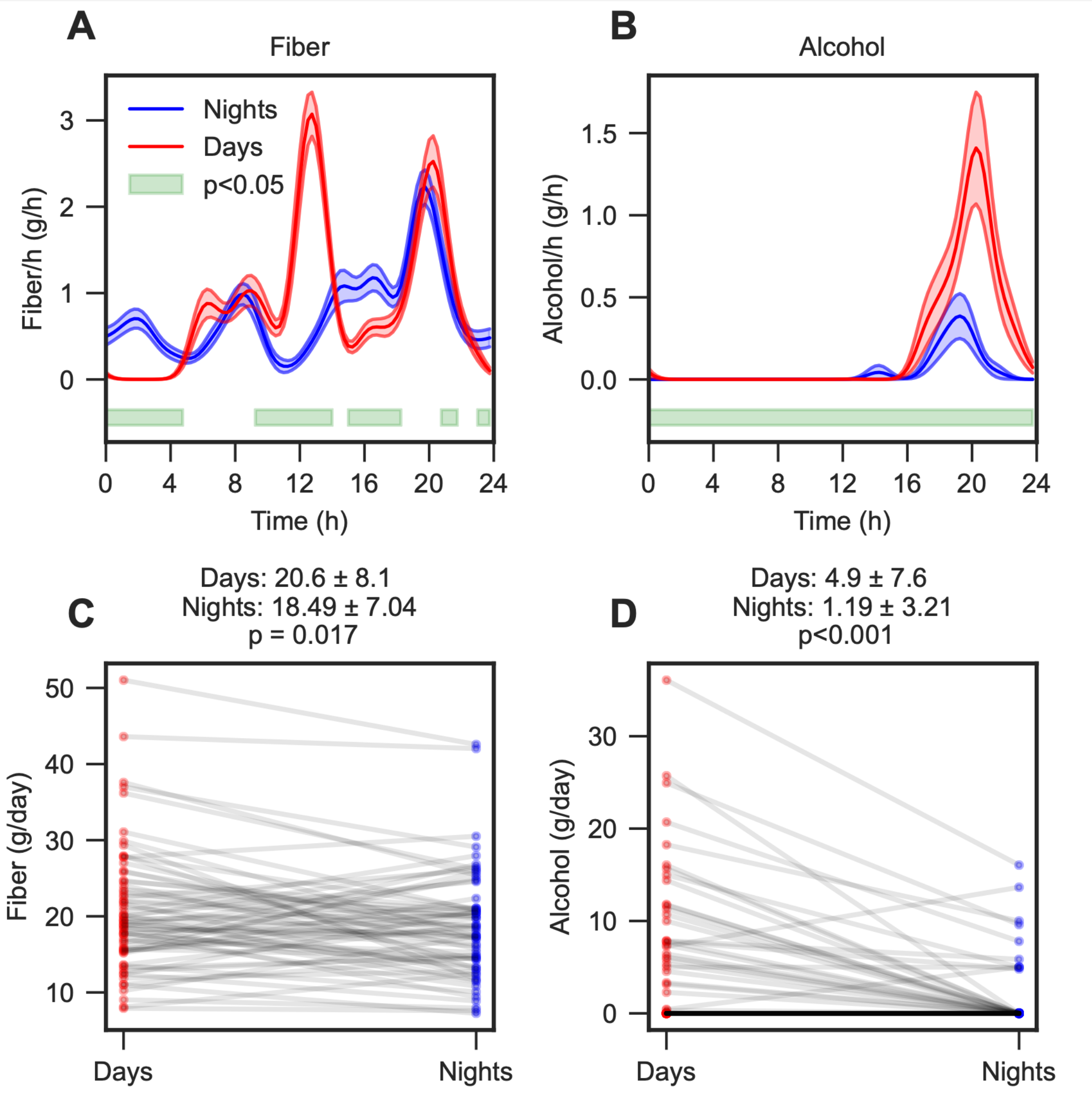


**Figure S2**. Temporal distribution of fiber and alcohol consumption during day and night shifts (*related to Figure 1*). (A-B) The mean consumption rate per hour for night shifts (blue) and day shifts (red) for fiber and alcohol, respectively. The shaded area represents the mean ± SEM, and the green bar represents significant group differences as assessed from $p<0.05$ in a Wilcoxon non-parametric test, adjusted for multiple testing with the Benjamini-Hochberg procedure. (C-D) Individual average daily intake for the day and night shift period for fiber and alcohol, respectively. Group means ± SD are indicated above each panel. $n = 66$ for all analyses.
